# Comparative whole genome analysis reveals re-emergence of typical human Wa-like and DS-1-like G3 rotaviruses after Rotarix vaccine introduction in Malawi

**DOI:** 10.1101/2022.10.04.22280678

**Authors:** Chimwemwe Mhango, Akuzike Banda, End Chinyama, Jonathan J. Mandolo, Orpha Kumwenda, Chikondi Malamba-Banda, Kayla G. Barnes, Benjamin Kumwenda, Kondwani Jambo, Celeste M. Donato, Mathew D. Esona, Peter N. Mwangi, A. Duncan Steele, Miren Iturriza-Gomara, Nigel A. Cunliffe, Valentine N. Ndze, Arox W. Kamng’ona, Francis E. Dennis, Martin M. Nyaga, Chrispin Chaguza, Khuzwayo C. Jere

## Abstract

Genotype G3 rotaviruses rank among the most common rotavirus strains worldwide in humans and animals. However, despite a robust long-term rotavirus surveillance system from 1997 in Blantyre, Malawi, these strains were only detected from 1997 to 1999 and then disappeared and re-emerged in 2017, five years after the introduction of the Rotarix rotavirus vaccine. Here we analysed 27 whole genome sequences to understand how G3 strains re-emerged in Malawi. We randomly selected samples each month between November 2017 and August 2019 from stool samples of children hospitalised with acute diarrhoea at the Queen Elizabeth Hospital in Blantyre, Malawi. We found three genotypes namely G3P[4] (*n*=20), G3P[6] (*n*=1) and G3P[8] (*n*=6) associated with the re-emergence of G3 strains in Malawi post-Rotarix vaccine introduction. The identified genotypes co-circulated at different time points and were associated with three typical human G3 strains consisting of either a Wa-like or DS-1-like genetic constellation and reassortant strains possessing Wa-like and DS-1-like genetic backbones. Time-resolved phylogenetic trees demonstrated that the most recent common ancestor for each segment of the re-emerged G3 strains emerged between 1996 and 2012, possibly through introductions from outside the country due to the limited genetic similarity with G3 strains which circulated before their disappearance in the late 1990s. Further genomic analysis revealed that the reassortant DS-1-like G3P[4] strains acquired a Wa-like NSP2 genome segment (N1 genotype) through intergenogroup reassortment; an artiodactyl-like VP3 through intragenogroup interspecies reassortment; and VP6, NSP1 and NSP4 segments through intragenogroup reassortment likely before importation into Malawi. Additionally, the re-emerged G3 strains contain amino acid substitutions within the antigenic regions of the VP4 proteins which could potentially impact the binding of rotavirus vaccine-induced antibodies. Altogether, our findings shows that multiple rather than a single genotype have driven the re-emergence of G3 strains likely from other countries highlighting the role of human mobility and genome reassortment events in the dissemination and evolution of rotavirus strains in Malawi necessitating the need for long-term genomic surveillance of rotavirus in high disease burden settings to inform disease prevention and control.

## Introduction

Childhood vaccination remains the most effective public health intervention against rotavirus gastroenteritis (Chandran et al. 2010). Despite the introduction of rotavirus vaccines in 114 countries globally (“VIEW-Hub by IVAC” n.d.), rotavirus remains the leading etiological agent of acute gastroenteritis in children (Clark et al. 2017), and is associated with approximately 128,500 deaths per annum among children <5 years old worldwide (Troeger et al. 2018). To reduce the global burden of gastroenteritis in children, the World Health Organisation (WHO) has pre-qualified the use of four rotavirus vaccines: Rotarix (GlaxoSmithKline, Rixensart, Belgium), RotaTeq (Merck and Co., Whitehouse Station, NJ, USA), ROTAVAC (Bharat Biotech, India) and ROTASIIL (Serum Institute of India Pvt. Ltd., India) (Kirkwood and Steele 2018). These vaccines are highly effective at preventing mortality, hospitalizations, and severe rotavirus gastroenteritis episodes in high-income settings, although lower effectiveness has been reported in low- and middle-income countries (Henschke et al. 2022; Cates, Tate, and Parashar 2022; Saha et al. 2021). The Rotarix vaccine was introduced into Malawi’s Extended Program of Immunisation (EPI) in October 2012, and by 2016, over 99% of vaccine coverage was reached (Bennett et al. 2021). The Rotarix vaccine has demonstrated relatively modest effectiveness in Malawi (31.7 to 70.6%) during programmatic use compared with that observed in high income settings, although effectiveness against severe rotavirus gastroenteritis in the first year of life is higher against homotypic (70%) compared with heterotypic (40 to 60%) strains (Bar-Zeev et al. 2016).

Rotavirus has a double-stranded RNA genome and belongs to the *Reoviridae* family. Its genome contains eleven segments that encode six structural proteins (VP1-VP4, VP6 and VP7) and up to six non-structural proteins (NSP1-NSP5/6) (Estes et al. 2007). To fully characterise rotavirus strains, a whole genome classification system was devised to assign genotypes to each genome segment (Matthijnssens, Ciarlet, Rahman, et al. 2008). To date, 41 G, 57 P, 31 I, 27 R, 23C, 23 M, 38 A, 27 N, 27 T, 31 E, and 27 H genotypes have been assigned for the VP7, VP4,VP6, VP1, VP2, VP3, NSP1, NSP2, NSP3, NSP4 and NSP5 genome segments, respectively (https://rega.kuleuven.be/cev/viralmetagenomics/virus-classification). While most infections are associated with a single genotype, coinfections with rotavirus strains of different genotypes are also common, which provides favourable conditions for generating progeny viruses with reassortant genomic segments. These genomic reassortment events together with high mutation rates arising from an error-prone RNA polymerase (VP1) that lacks proof-reading mechanisms are the main mechanisms of evolution for rotaviruses (Estes et al. 2007).

Clinically, genotypes G1P[8], G2P[4], G3P[8], G4P[8], G9P[8] and G12P[8] are the most common rotavirus strains associated with diarrhoea in under five children globally (Dóró et al. 2014). The distribution of these and other rotavirus genotypes vary by geographical location and appear to depend on the rotavirus vaccination status and the vaccine used in a particular region (Donato et al. 2022). Before global introduction of rotavirus vaccines across the continents, G1P[8] genotypes were the predominant strains detected globally (Bányai et al. 2012). However, G2P[4] strains have been predominant in some countries using Rotarix rotavirus vaccine while the G12P[8] genotype has been commonly detected in some countries using RotaTeq rotavirus vaccine (Roczo-Farkas et al. 2018; Hungerford et al. 2019; Carvalho-Costa et al. 2019; Vizzi et al. 2017; Degiuseppe and Stupka 2018). The detection rates of G3 strains and other sporadically circulating genotypes has increased especially in countries using Rotarix rotavirus vaccine globally (Carvalho-Costa et al. 2019; Wahyuni et al. 2021; Roczo-Farkas et al. 2018). While genotype G3 was not common in the African continent before rotavirus vaccine introduction, these strains have been reported in several African countries post-vaccination (Mhango et al. 2020; Mwanga et al. 2020; João et al. 2020). Our previous work reported the prevalence of rotavirus strains circulating in Blantyre, Malawi from 1997 to 2019 (Mhango et al. 2020). We showed that genotypes G1, G4 and G8 were frequently detected before Rotarix rotavirus vaccine introduction, whereas G1, G2, G3 and G12 genotypes were more common during the vaccine era (Mhango et al. 2020). Although G3 strains were last detected between 1997 to 1999 before their re-emergence in 2017 after nearly two decades, they became the most predominant rotavirus strains in Blantyre, Malawi by the end of 2019 following their re-emergence in 2017 (Mhango et al. 2020).

Although G3 rotaviruses are associated with a wide range of host species and “P” genotype combinations (Martínez-Laso et al. 2009; Matthijnssens et al. 2006), it’s unknown whether the re-emerged G3 rotaviruses belong to a single genotype, particularly P[8] which constitutes the majority of human-associated strains (Matthijnssens, Ciarlet, Heiman, et al. 2008). It’s also unknown whether the genetic background of the re-emerged G3 strains in Malawi possess a typical human or equine-like VP7 genome segment and DS-1-like genetic backbone as recently seen in various settings globally after the introduction of rotavirus vaccines (Komoto et al. 2018; Cowley et al. 2016; Dóró et al. 2016; Esposito et al. 2019; Arana et al. 2016; Luchs et al. 2019). To address these questions, we generated whole genome sequences of G3 strains collected through our robust and long-term hospital-based rotavirus surveillance in Blantyre, Malawi to investigate their genomic epidemiology and evolution in Malawi and broader international context. Our findings show that re-emergence of G3 rotavirus strains was driven by multiple genotypes, including reassortant strains, with highest genetic similarity with strains from other countries, highlighting the impact of importation events as a mechanism for reseeding the strains in the post-vaccination era following their nearly two decades hiatus in Malawi.

## Results

### G3 rotaviruses strains became the predominant genotype in Malawi after their re-emergence in November 2017

Active rotavirus surveillance has been on-going at QECH from 1997 of which a wide diversity of rotavirus strains has been reported from stools collected from children presenting with diarrhoea during weekdays (Mhango et al. 2020). G3 strains were last detected in 1999 (0.02% of total genotypes; 4/166) and re-emerged in November 2017, five years after the introduction of the rotavirus vaccine. By 2018, G3 strains had completely replaced G1 and G2 strains as the most commonly detected rotavirus strains in Malawi (Mhango et al. 2020). Genotype G3P[4] strains re-emerged first in 2017 and were detected at high frequencies (12.3 to 36.3% of all genotypes), G3P[6] strains were sporadically detected (1.2%) between September 2018 to March 2019 where as G3P[8] were first detected in December 2018 and predominated in 2019 (27.2% of all genotypes) (Mhango et al. 2020). To gain insight into the evolutionary events that led to the re-emergence of G3 rotaviruses in Malawi, full length nucleotide sequences for 30 G3 strains that circulated before and after vaccine introduction periods were generated and analysed (Figure 1 and Supplementary Table S1).

**Figure 1.**
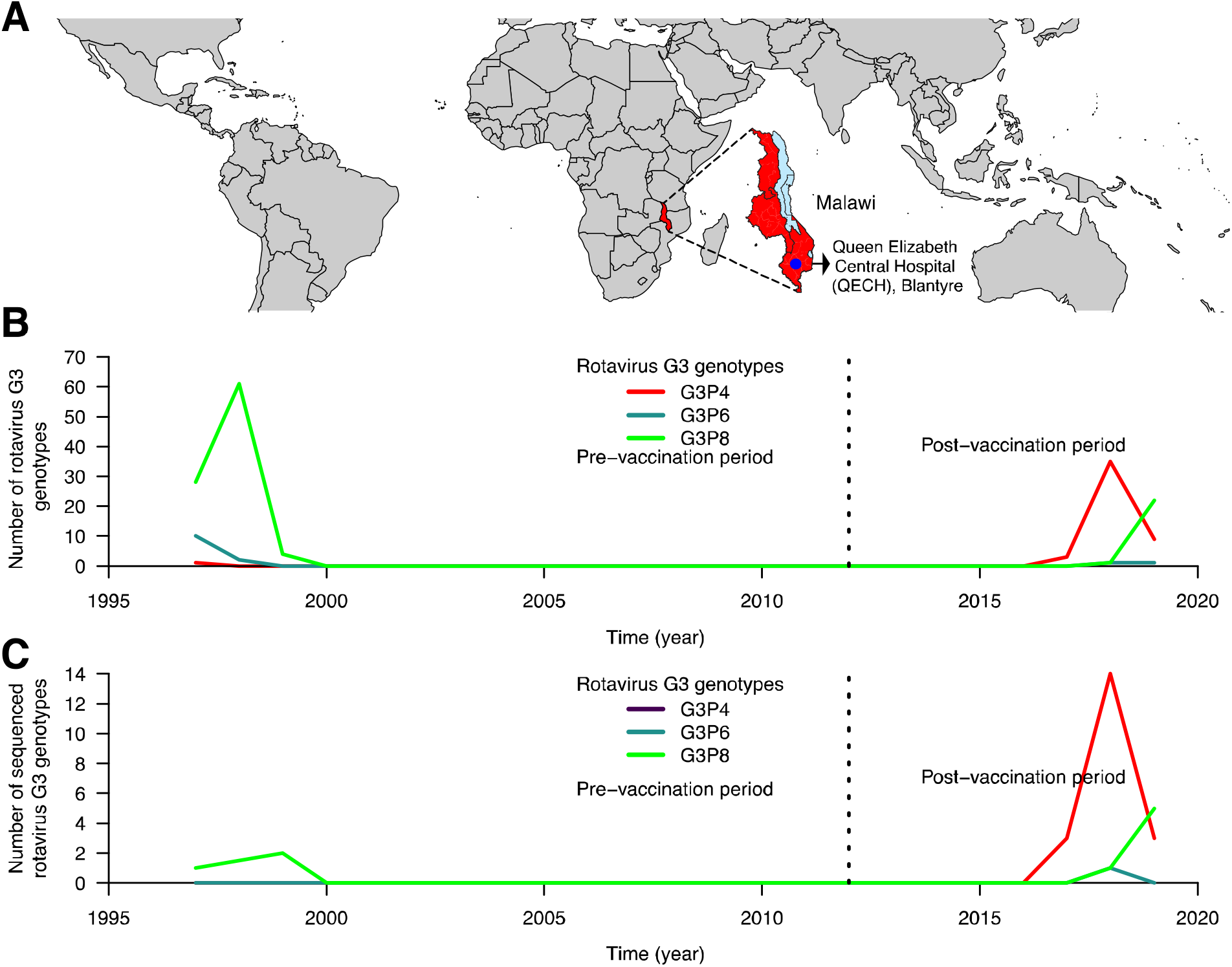
Rotavirus G3 genotypes and whole genome sequences characterised at Queen Elizabeth Central Hospital, Malawi before and after rotavirus vaccine introduction. **(A)** Global map highlighting the geographical location of Malawi in Africa as well as the location of Queen Elizabeth Central Hospital (QECH) in Malawi where the diarrhoea surveillance platform is set up. **(B)** Absolute number of PCR positive G3 rotavirus strains genotyped before and after Rotarix rotavirus vaccine introduction at QECH. (C) Absolute number of G3 whole genome sequences generated from stool specimens collected before and after Rotarix rotavirus vaccine introduction in Malawi.

### Re-emergent G3 rotaviruses in Malawi were associated with four variants

G3 strains are associated with a wider host range which predisposes them to reassortment events as well as emergence of unusual G3 variants (Martínez-Laso et al. 2009; Matthijnssens et al. 2006). Normally, the inner capsid genome segments of human rotaviruses have either a Wa-like (I1-R1-C1-M1-A1-N1-T1-E1-H1), DS-1-like (I2-R2-C2-M2-A2-N2-T2-E2-H2) or AU-1-like (I3-R3-C3-M3-A3-N3-T3-E3-H3) genotype constellation. To gain insights in the genotype constellations of the G3 variants that re-emerged and predominated between 2017 to 2019 in Blantyre, Malawi, we analysed the whole genome sequences (WGS) of 27 representative G3 strains that re-emerged to determine the genomic constellations of the viruses. WGS analysis revealed that sixteen G3P[4] (80%, *n=*20) strains had a pure DS-1-like genotype constellation (Table 1). The only G3P[6] strain that was sequenced also had a pure DS-1-like genotype constellation (Table 1). In contrast, the three G3P[8] strains detected from 1997 to 1999 (33.3%, *n*=9) before rotavirus vaccine introduction and six from 2018 to 2019 (66.6%, *n*=9) during the rotavirus vaccine era in Malawi had the Wa-like genotype constellation (Table 1). Unexpectedly, four G3P[4] (20%, *n*=20) strains that were detected towards the end of 2018 had a mosaic genotype constellation consisting of a core DS-1-like backbone but with an N1 NSP2 Wa-like rather than an N2 DS-1-like genome segment. We therefore classified these atypical viruses as reassortant DS-1-like G3P[4] strains (Table 1). Thus, a total of four genotype G3 variants circulated between 2017 and 2019, namely the typical DS-1-like strains (G3P[4] and G3P[6]), reassortant DS-1-like strains (G3P[4]) and Wa-like strains (G3P[8]), of which the later two co-circulated in late 2018.

**Table 1.** Whole genotype constellations of pre- and post-vaccine Malawian G3 strains. The nomenclature of all the rotavirus strains indicates the rotavirus group, species where the strain was isolated, name of the country where the strain was originally isolated, the common name, year of isolation and the genotypes for genome segment 4 and 9 as proposed by the *Rotavirus Classification Working Group* (RCWG) (Matthijnssens et al. 2008).

| Strain Nomenclature | Genotype constellation |  |  |  |  |  |  |  |  |  |  | Genogroup/<br>Constellation |
| --- | --- | --- | --- | --- | --- | --- | --- | --- | --- | --- | --- | --- |
| Pre-vaccine strains | VP7 | VP4 | VP6 | VP1 | VP2 | VP3 | NSP1 | NSP2 | NSP3 | NSP4 | NSP5 |  |
| RVA/Human-wt/MWI/MW1-001/1997/G3P[8] | G3 | P[8] | I1 | R1 | C1 | M1 | A1 | N1 | T1 | E1 | H1 | Wa-like |
| RVA/Human-wt/MWI/MW1-621/1999/G3P[8] | G3 | P[8] | I1 | R1 | C1 | M1 | A1 | N1 | T1 | E1 | H1 | Wa-like |
| RVA/Human-wt/MWI/OP1-511/1999/G3P[8] | G3 | P[8] | I1 | R1 | C1 | M1 | A1 | N1 | T1 | E1 | H1 | Wa-like |
| Post-vaccine strains |  |  |  |  |  |  |  |  |  |  |  |  |
| RVA/Human-wt/MWI/BTY22H/2017/G3P[4] | G3 | P[4] | I2 | R2 | C2 | M2 | A2 | N2 | T2 | - | H2 | DS-1-like |
| RVA/Human-wt/MWI/BTY22J/2017/G3P[4] | G3 | P[4] | I2 | R2 | C2 | M2 | A2 | N2 | T2 | E2 | H2 | DS-1-like |
| RVA/Human-wt/MWI/BTY232/2017/G3P[4] | G3 | P[4] | I2 | R2 | C2 | M2 | A2 | N2 | T2 | E2 | H2 | DS-1-like |
| RVA/Human-wt/MWI/BTY23I/2018/G3P[4] | G3 | P[4] | I2 | R2 | C2 | M2 | A2 | N2 | T2 | E2 | - | DS-1-like |
| RVA/Human-wt/MWI/BTY240/2018/G3P[4] | G3 | P[4] | I2 | R2 | C2 | M2 | A2 | N2 | T2 | E2 | H2 | DS-1-like |
| RVA/Human-wt/MWI/BTY24R/2018/G3P[4] | G3 | P[4] | I2 | R2 | C2 | M2 | A2 | N2 | T2 | E2 | H2 | DS-1-like |
| RVA/Human-wt/MWI/BTY25Q/2018/G3P[4] | G3 | P[4] | I2 | R2 | C2 | M2 | A2 | N2 | T2 | E2 | H2 | DS-1-like |
| RVA/Human-wt/MWI/BTY25L/2018/G3P[4] | G3 | P[4] | I2 | R2 | C2 | M2 | A2 | N2 | T2 | E2 | H2 | DS-1-like |
| RVA/Human-wt/MWI/BTY26Q/2018/G3P[4] | G3 | P[4] | I2 | R2 | C2 | M2 | A2 | N2 | T2 | E2 | H2 | DS-1-like |
| RVA/Human-wt/MWI/BTY26H/2018/G3P[4] | G3 | P[4] | I2 | R2 | C2 | M2 | A2 | N2 | T2 | E2 | H2 | DS-1-like |
| RVA/Human-wt/MWI/BTY27C/2018/G3P[4] | G3 | P[4] | I2 | R2 | C2 | M2 | A2 | N2 | T2 | E2 | H2 | DS-1-like |
| RVA/Human-wt/MWI/BTY27S/2018/G3P[4] | G3 | P[4] | I2 | R2 | C2 | M2 | A2 | N2 | T2 | E2 | H2 | DS-1-like |
| RVA/Human-wt/MWI/BTY296/2018/G3P[4] | G3 | P[4] | I2 | R2 | C2 | M2 | A2 | N2 | T2 | E2 | H2 | DS-1-like |
| RVA/Human-wt/MWI/BTY29D/2018/G3P[4] | G3 | P[4] | I2 | R2 | C2 | M2 | A2 | N2 | T2 | E2 | H2 | DS-1-like |
| RVA/Human-wt/MWI/BTY29E/2018/G3P[4] | G3 | P[4] | I2 | R2 | C2 | M2 | A2 | N2 | T2 | E2 | - | DS-1-like |
| RVA/Human-wt/MWI/BTY2A2/2018/G3P[4] | G3 | - | I2 | R2 | C2 | M2 | A2 | N2 | T2 | E2 | - | DS-1-like |
| RVA/Human-wt/MWI/BTY2CM/2018/G3P[6] | G3 | P[6] | I2 | R2 | C2 | M2 | A2 | N2 | T2 | E2 | H2 | DS-1-like |
| RVA/Human-wt/MWI/BTY2BG/2018/G3P[4] | G3 | P[4] | I2 | R2 | C2 | M2 | A2 | N1 | T2 | E2 | H2 | Reassortant |
| RVA/Human-wt/MWI/BTY2EP/2019/G3P[4] | G3 | P[4] | I2 | R2 | C2 | M2 | A2 | N1 | T2 | E2 | - | Reassortant |
| RVA/Human-wt/MWI/CHX11Q/2019/G3P[4] | G3 | P[4] | I2 | R2 | C2 | M2 | A2 | N1 | T2 | E2 | H2 | Reassortant |
| RVA/Human-wt/MWI/CHX11X/2019/G3P[4] | G3 | P[4] | I2 | R2 | C2 | M2 | A2 | N1 | T2 | E2 | - | Reassortant |
| RVA/Human-wt/MWI/BTY2BD/2018/G3P[8] | G3 | P[8] | I1 | R1 | C1 | M1 | A1 | N1 | T1 | E1 | H1 | Wa-like |
| RVA/Human-wt/MWI/BTY2EM/2019/G3P[8] | G3 | P[8] | I1 | R1 | C1 | M1 | A1 | N1 | T1 | E1 | H1 | Wa-like |
| RVA/Human-wt/MWI/BTY2GA/2019/G3P[8] | G3 | P[8] | I1 | R1 | C1 | M1 | A1 | N1 | T1 | E1 | H1 | Wa-like |
| RVA/Human-wt/MWI/BTY2GC/2019/G3P[8] | G3 | P[8] | I1 | R1 | C1 | M1 | A1 | N1 | T1 | E1 | H1 | Wa-like |
| RVA/Human-wt/MWI/CHX11U/2019/G3P[8] | G3 | P[8] | I1 | R1 | C1 | M1 | A1 | N1 | T1 | E1 | H1 | Wa-like |
| RVA/Human-wt/MWI/CHX11S/2019/G3P[8] | G3 | P[8] | I1 | R1 | C1 | M1 | A1 | N1 | T1 | E1 | H1 | Wa-like |

Genome segments of segmented viruses tend to have evolutionary rates that vary per segment. This coupled with reassortment events tend to increase genomic diversity of segmented viruses. To assess the genomic diversity within the re-emergent G3 variants, we placed the VP7, VP4 and inner capsid genome segments for DS-1-like G3 variants into global context based on the lineages defined by Sadiq et al. and Agbemabiese et al. (Agbemabiese et al. 2019; Sadiq et al. 2019). We could not do the same for the inner capsid segments of the Wa-like strains since there is no known lineage framework for Wa-like genome segments. WGS analysis revealed a wider genomic diversity (1 to 4) per genome segment for the re-emerged four G3 variants (Supplementary Table S2). The highest diversity was observed within VP4 while the lowest was seen in VP7 (Supplementary Table S2). However, we also observed within lineage variation for DS-1-like variants (VP1, VP2, VP3, and NSP3) having up to 2, while NSP5 had up to three unique clusters within the same lineage (Supplementary Figure S3). Although we only identified four variants associated with the G3 rotavirus strains, the association of the G3 strains with multiple lineages suggest a high genomic diversity of the re-emerged G3 strains circulated in Blantyre, Malawi between 2017 and 2019.

### Re-emergent G3 strains were genetically distinct from previous and co-circulating rotaviruses in Malawi

Naturally, reassortment events are common within rotaviruses when multiple strains infect a single host. Since our present findings revealed a co-circulation of Wa-like and DS-1-like G3 strains late in 2018; we therefore explored the possibility that the reassortant DS-1-like G3P[4] strains acquired a Wa-like NSP2 genome segment through coinfections of Wa-like and DS-1-like strains that co-circulated in Malawi. We constructed a Maximum likelihood phylogenetic tree for all Malawian Wa-like NSP2 (N1) genome segment to explore their genetic relationship to the NSP2 of the reassortant DS-1-like G3P[4] strains. The phylogenetic analysis revealed that the Wa-like NSP2 genome segment (N1 genotype) of the reassortant DS-1-like G3P[4] strains formed a separate monophyletic cluster from the N1 NSP2 genome segments that were associated with co-circulating Wa-like G3P[8] and other locally detected non-G3 strains (Supplementary Figure S2). The NSP2 genome segment of the reassortant DS-1-like G3P[4] and other co-circulating Wa-like Malawian strains differed by 20-23 SNPs within the NSP2 ORF sequence. These findings suggested that the NSP2 genome segment of the re-emerged G3 strains was not acquired from strains circulating in Malawi, especially after the re-emergence of the G3, as these could have most likely been detected by our rotavirus surveillance system.

Previous WGS work in Malawi revealed a circulation of non-G3 genotypes on either a Wa-like or a DS-1-like genomic constellation (Jere et al. 2018). Considering the segmented nature of rotaviruses, we conducted a genome segment specific time resolved phylogenetic analysis to assess the genomic similarity of the re-emerged strains to locally detected co-circulating non-G3 Wa-like or DS-1-like rotaviruses in Malawi. When we employed the time tree analysis to compare the nucleotide sequences of the VP4 and nine backbone genome segments, we observed that the pre- and post-vaccine G3 strains formed separate monophyletic clades from the co-circulating non-G3 Wa-like and DS-1-like strains (Figure 2 and Supplementary Figure S1). These findings suggested that the re-emerged G3 strains did not evolve from or acquire their genome segments from the other co-circulating non-G3 rotaviruses raising the possibility of new introduction of these strains into Malawi.

**Figure 2.**
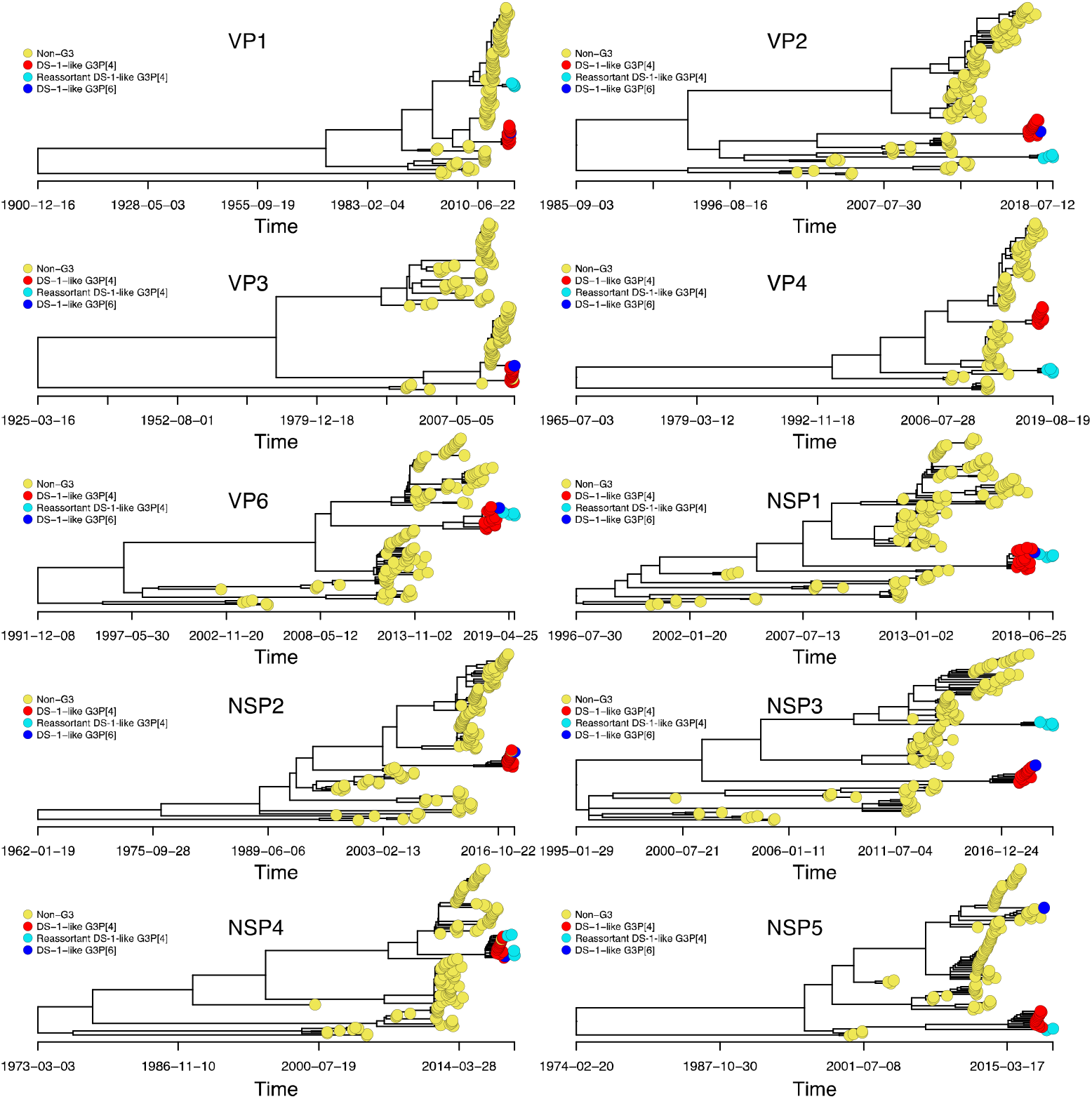
Time resolved phylogenetic trees for DS-1-like genome segments associated with G3 as well as non-G3 rotavirus strains detected in Malawi from 1997 to 2019. Only previously circulating DS-1-like genome segments with a complete open reading frame were used to estimate the time to the most common recent ancestor for the DS-1-like (G3P[4], G3P[6] and reassortant G3P[4]) in relation to locally circulating strains. Time trees were constructed using Nextstrain. The VP4 tree only contains genotype P[4]. The tips of DS-1-like G3P[4], DS-1-like G3P[6] and reassortant DS-1-like G3P[4] segments are annotated in red, blue and light blue colours respectively while all non-G3 genome segments are annotated in yellow colours.

When we focused on the clustering patterns of the G3 strains, we identified two distinct populations for genome segments encoding both structural and non-structural proteins for Malawian G3P[4] strains that formed separate monophyletic clades. The monophyletic clades for the DS-1-like G3P[4] strains clustered away from the reassortant DS-1-like G3P[4] strains in VP1, VP2, VP4, NSP3 and NSP5 encoding genome segments (Figure 2). Even though the reassortant DS-1-like G3P[4] strains emerged after the DS-1-like G3P[4] and shared similar NSP1, NSP4 and VP6 genome segments, the tMRCA of the NSP3, NSP5, VP1, VP2 and VP4 genome segments for the reassortant DS-1-like G3P[4] strains (ranging from 2001 – 2010) (Table 2) and that of the DS-1-like G3P[4] strains (ranging from 2001 – 2008) (Table 2) varied considerably across the respective segments. When we looked at the estimated mutation rates for each of the genome segments (NSP3, NSP5, VP1, VP2, VP3 and VP4) (2.52 × 10^−4^ to 1.67 × 10^−3^ nucleotide substitutions per site per year) (Table 2), the number of single nucleotide polymorphisms (SNPs) and the time between the initial emergence of the two G3P[4] variants, we concluded that there was not enough time for these specific genome segments of reassortant DS-1-like G3P[4] to have evolved from the DS-1-like genome segments. Thus, these data suggested that the DS-1-like G3P[4] and reassortant DS-1-like G3P[4] strains did not share an immediate common ancestor and that the reassortant DS-1-like G3P[4] strains were introduced independently already having a reassortant NSP2 genome segment from a source where DS-1-like G3P[4] strains similar to those detected in Blantyre, Malawi during this period were also circulating.

**Table 2.** Time to the most recent common ancestor (tMRCA) for Malawian Wa-like and DS-1-like G3 strains and estimated mutation rates for Wa-like and DS-1-like time trees. The time to the most common recent ancestors (tMRCA) for G3 strains were estimated in relation to locally circulating human DS-1-like and Wa-like genome segments. The tMRCA and mutation rates were extracted from the time trees generated in Nextstrain for Figure 2 and Supplementary Figure S1.

| Genome Segment<br>(Protein) | Wa-like G3P[8] |  | DS-1-like G3P[4] |  | Reassortant DS-1-like G3P[4] |
| --- | --- | --- | --- | --- | --- |
|  | tMCRA (95% CI) | Mutation rates* | tMCRA (95% CI) | Mutation rates | tMCRA (95% CI) |
| **1 (VP1)** | 21 May 1996<br>(25 Jul 1990 - 20 Sep 1998) | 8.15 x 10<sup>-4</sup> | 11 Feb 2008<br>(21 Jul 2006 - 10 May 2009) | 8.09 x 10<sup>-4</sup> | 12 Feb 2008<br>(30 Dec 2005 - 15 Feb 2009) |
| **2 (VP2)** | 03 Jan 2009<br>(06 Aug 2007 - 06 May 2010) | 9.35 x 10<sup>-4</sup> | 18 Oct 2002<br>(21 Mar 2000 - 15 Mar 2005) | 6.43 x 10<sup>-4</sup> | 17 Oct 2002<br>(24 Nov 1999 - 03 Sep 2004) |
| **3 (VP3) a** | 12 Jul 2000<br>(08 Oct 1999 - 11 Nov 2001) | 1.04 x 10<sup>-3</sup> | 05 Jan 2012<br>(01 Apr 2010 - 22 May 2012) | 1.67 x 10<sup>-3</sup> | - |
| **4 (VP4)** | 03 Dec 2008<br>(14 Nov 2007 - 29 Mar 2010) | 8.31 x 10<sup>-4</sup> | 07 Sept 2007<br>(13 Oct 2003 - 16 Nov 2010) | 9.14 x 10<sup>-4</sup> | 28 May 2010<br>(16 Jul 2006 - 20 Aug 2013) |
| **6 (VP6)** | 26 Nov 2006<br>(28 Jun 2003 - 24 Nov 2013) | 6.37 x 10<sup>-4</sup> | 14 Dec 2008<br>(20 Oct 2002 - 18 Nov 2013) | 8.30 x 10<sup>-4</sup> | 14 Dec 2008<br>(20 Oct 2002 - 18 Nov 2013) |
| **5 (NSP1)** | 15 Mar 2001<br>(21 Jul 2000 - 17 Jan 2004) | $1.55 \times 10^{-3}$ | 15 Jul 2007<br>(10 Sept 2004 - 17 Jul 2009) | $1.44 \times 10^{-3}$ | 15 Jul 2007<br>(10 Sept 2004 - 17 Jul 2009) |
| **7 (NSP2) b** | - | - | 17 Jan 2003<br>(08 Jan 1996 - 28 Jan 2005) | $6.59 \times 10^{-4}$ | - |
| **8 (NSP3)** | 16 Oct 2005<br>(10 Jun 2003 - 31 May 2008) | $4.23 \times 10^{-4}$ | 21 Jul 2001<br>(29 Aug 1997 - 10 May 2006) | $7.55 \times 10^{-4}$ | 11 May 2009<br>(13 Jan 2006 - 24 May 2012) |
| **10 (NSP4)** | 29 Mar 2003<br>(16 Aug 2000 - 12 Jun 2004) | $8.46 \times 10^{-4}$ | 25 Feb 2010<br>(15 Jul 2005 - 09 Jan 2012) | $1.43 \times 10^{-3}$ | 25 Feb 2010<br>(15 Jul 2005 - 09 Jan 2012) |
| **11 (NSP5)** | 06 Jan 1995<br>(19 Nov 1984 - 24 Jun 2001) | $2.52 \times 10^{-4}$ | 21 May 2001<br>(05 Feb 1998 - 25 Jan 2005) | $6.09 \times 10^{-4}$ | 21 May 2001<br>(05 Feb 1998 - 25 Jan 2005) |

**a.** tMRCA were not estimated for reassortant DS-1-like G3P[4] genome segments because they resembled animal-like genome segments thus we could not estimate the correct tMRCA using the current dataset.
**b.** tMRCA were not estimated for Wa-like NSP2 genome segments due to a lack of a sufficient molecular clock signal to conduct the analysis.
\*Mutation rates were defined as the number of substitutions per site per year.

### Re-emerged G3 strains resembled typical human rotaviruses likely imported into Malawi

Human G3 rotaviruses have been detected at higher frequencies globally during the past decade with the majority possessing equine-like rotavirus genome segments. While the present study G3 strains had either a DS-1-like or Wa-like genomic constellation, the origins of the genome segments were unclear. To determine the potential host origins of the re-emerged G3 strains, we performed a maximum likelihood phylogenetic analysis for the VP7 genome segment of the Malawian and globally detected G3 strains (2010 - 2020). Maximum likelihood analysis revealed that the genome segments encoding VP7 for all Malawian G3 strains clustered together with those of the typical human G3 strains that were detected from various countries across the globe (Figure 3). Except for the VP3, the rest of the genome segments clustered together with genome segments characterised from other human rotaviruses (Supplementary Figure S3). The VP3 (M2) genome segments of the reassortant G3P[4] strains clustered together with M2 genome segments commonly characterised in ruminant animals (Figure 4b). This suggested that intragenogroup reassortment events between human rotaviruses and strains that circulate in Artiodactyla order were part of the evolutionary events that led to the emergence of the reassortant DS-1-like G3P[4] strains.

**Figure 3.**
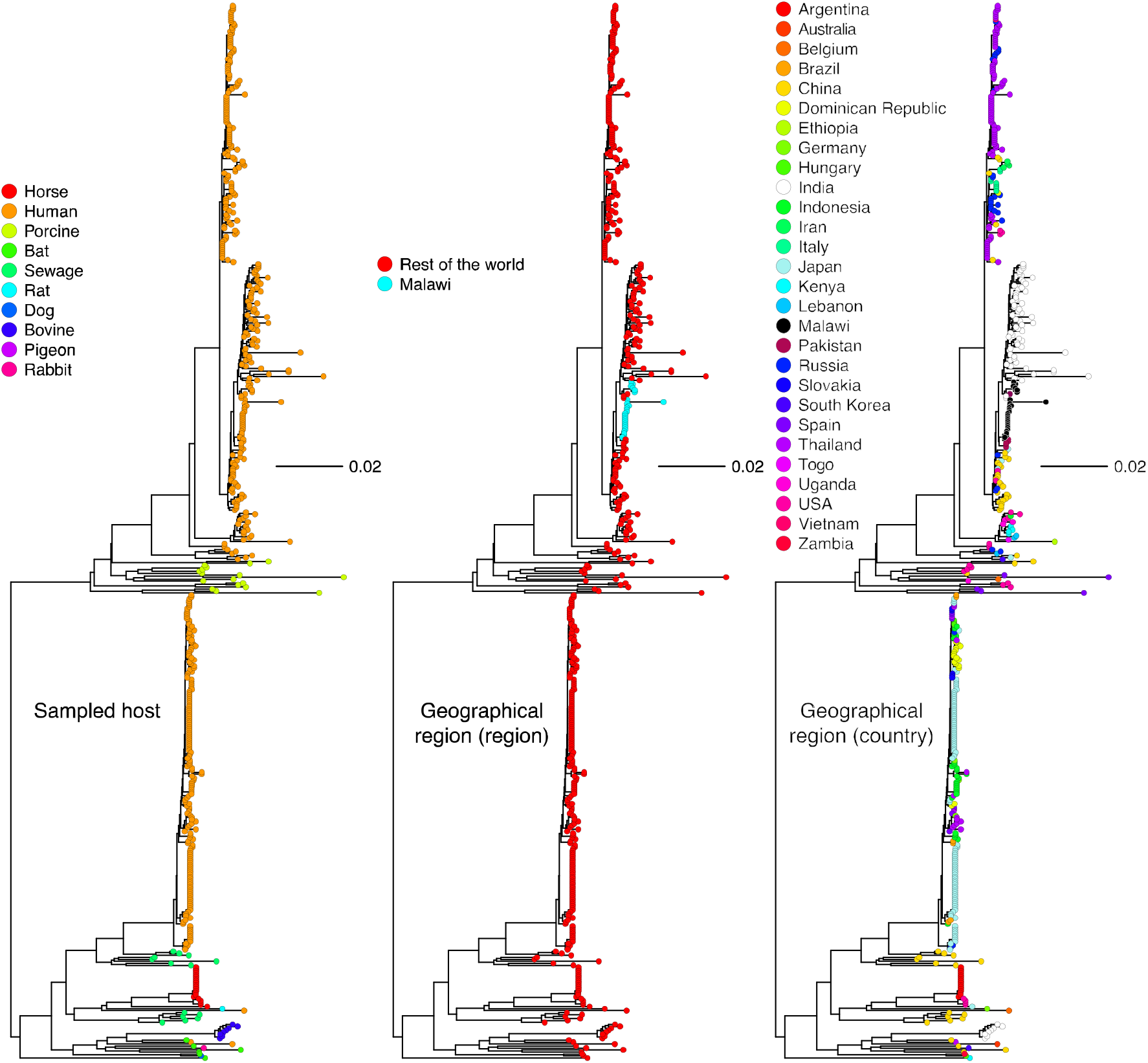
Global Maximum Likelihood phylogenetic trees for VP7 genome segments encoding the genotype G3. Only genome segments with a complete open reading frame characterised between 2010 to 2019 were included in the analysis. The GTR evolutionary model with Gamma heterogeneity across nucleotide sites was used for phylogenetic inference while 1000 botastraps were used to assess the reliability of the branching order. The tree was rooted using the RVA/Pigeon-wt/JPN/PO-13/1989/G18P[17] but the outgroups were omitted for better visualisation.

**Figure 4.**
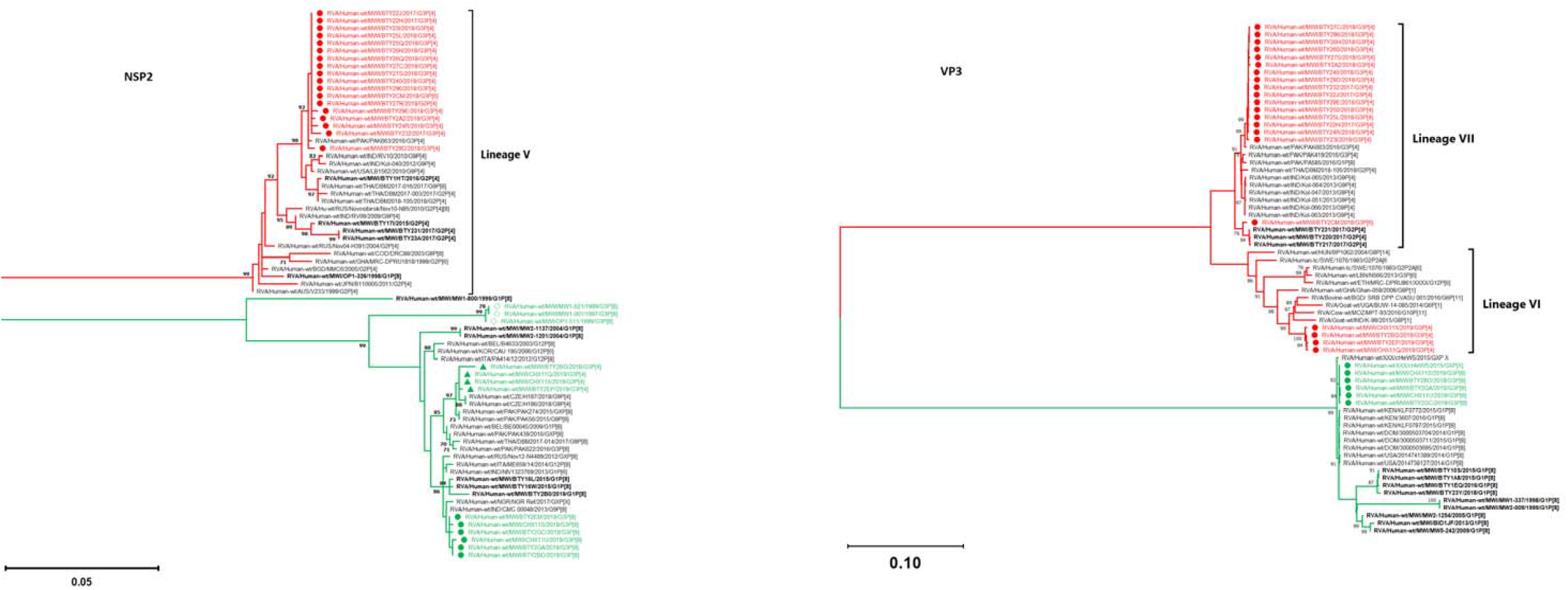
Maximum likelihood (ML) phylogenetic trees based on complete open reading frame of rotavirus NSP2 and VP3 genome segments. The trees were out grouped at RVA/Pigeon-wt/JPN/PO-13/1983/G18P[17] but were omitted for clear visualisation. The GTR evolutionary model with Gamma heterogeneity across nucleotide sites was used for phylogenetic inference. Bootstrap values ≥70% are shown adjacent to each branch node. Genotype 1 (Wa-like) and 2 (DS-1-like) branches are annotated in green and red respectively. The strain names of Malawian Wa-like and DS-1-like G3 genome segments are denoted by green and red colours respectively. Circles represent post-vaccine while diamonds represent pre-vaccine Malawian G3 strains. Reassortant G3P[4] are represented by green triangles in (NSP2).

We then looked at the geographical origin of the genome segments associated with G3 rotaviruses circulating in Malawi. Maximum likelihood analysis revealed that all VP7 genome segments associated with strains possessing a DS-1-like genetic backbone (G3P[4], reassortant G3P[4] and G3P[6]) shared a high nucleotide sequence similarity with only 4 to 6 SNPs observed within the ORF to contemporary G3P[4] strains characterised in the middle east Asia, Pakistan (Figure 3 and Supplementary Figure S3). In contrast, the VP7 genome segment of Wa-like (G3P[8]) strains clustered together with and only had 5 to 7 SNPs within the ORF to contemporary G3P[8] strains characterised in the far east Asia (Japan and Thailand) (Figure 3 and Supplementary Figure S3). The rest of the DS-1-like G3P[4] genome segments clustered together with isolates from the Asian continent with a highest nucleotide sequence similarity (99.40% to 99.70%) observed against contemporary G3P[4] strains that were detected in Pakistan (Figure 4 and Supplementary Figure S3). With the exception of NSP1 and VP4, the genome segments for the DS-1-like G3P[6] study strain had a similar clustering pattern to our study DS-1-like G3P[4] strains which showed a high nucleotide sequence similarity to African (Mozambique, Zambia, Zimbabwe and South Africa) DS-1-like isolates (Supplementary Figure S3). To the contrary, we observed two clustering patterns of the re-emerged reassortant DS-1-like G3P[4] strains in Malawi depending on the genome segment. While NSP1, NSP4 and VP6 showed a high nucleotide sequence identity to Asian (Pakistan) isolates, similar to the DS-1-like G3P[4] strains, their NSP2, NSP5, VP1, VP2 and VP4 showed a high nucleotide sequence identity (99.51% to 99.70%) to similar genome segments characterised in Europe (Czech Republic) (Figure 4, and Supplementary Figure S3). On the other hand, the Wa-like G3P[8] genome segments showed similarity to Wa-like segments for non-G3 strains characterised from the African (Zimbabwe, South Africa, Mozambique, Kenya and Rwanda) and Wa-like G3P[8] from the Asian (Japan, India and Indonesia) continents (Figure 4 and Supplementary Figure S3.). Although the majority of these findings showed a high similarity to Asian countries, these genome segments are widespread across the African, Asian, European, and potentially other unsampled settings suggesting a potential for frequent cross border dissemination.

### Reassortant DS-1-like G3P[4] likely emerged through multiple reassortment events prior to their introduction into Malawi

As all genome segments for the reassortant DS-1-like G3P[4] did not cluster with those of the other rotavirus strains that were circulating in Malawi, we used phylogenetic inferences for each genome segment to determine the origin of the reassortant strains. Since the VP1, VP2, VP4, NSP3 and NSP5 encoding genome segments of the reassortant DS-1-like G3P[4] were not closely related to currently available strains in the GenBank, presumably due to dearth of genomic rotavirus surveillance in many countries, we hypothesised that the reassortant DS-1-like G3P[4] acquired these five genome segments from human rotaviruses that circulated from unsampled locations. Our reassortant strains likely acquired their VP3 genome segment from artiodactyl rotaviruses through intragenogroup reassortment as the closest related cognate VP3 encoding genome segments were those of bovine strain MPT-93 detected in Mozambique in 2015 and a caprine strain K-98 detected in India in 2015. They likely acquired their NSP2 genome segment from Wa-like rotaviruses intergenogroup reassortment as it was assigned an N1 genotype. Phylogenetically, the NSP2 for our reassortant strains was closely related to those that were detected in the Czech Republic around 2019. We could not infer when and where exactly the NSP2 reassortment events occurred due to the limited numbers of available rotavirus whole genome sequence data from many countries. Similarly, the reassortant DS-1-like G3P[4] likely acquired or donated their DS-1-like VP6, VP7, NSP1 and NSP4 from G3P[4] rotaviruses that circulated in Islamabad and Rawalpindi in Pakistan from 2014-2016 (Umair et al. 2018; Sadiq et al. 2019; Naqvi et al. 2020) As limited countries are conducting rotavirus genomic surveillance, it is possible that these reassortment events occurred in an unsampled location prior to their importation into Pakistan. Nevertheless, the resultant reassortant DS-1-like G3P[4] strains were the ones that likely ended up in Malawi where they were associated with diarrhoea infections in 2017 and 2018 at QECH (Mhango et al. 2020) (Figure 5). Thus, the reassortant DS-1-like G3P[4] strains were likely generated through a series of reassortment events elsewhere prior to their circulation in Malawi.

**Figure 5.**
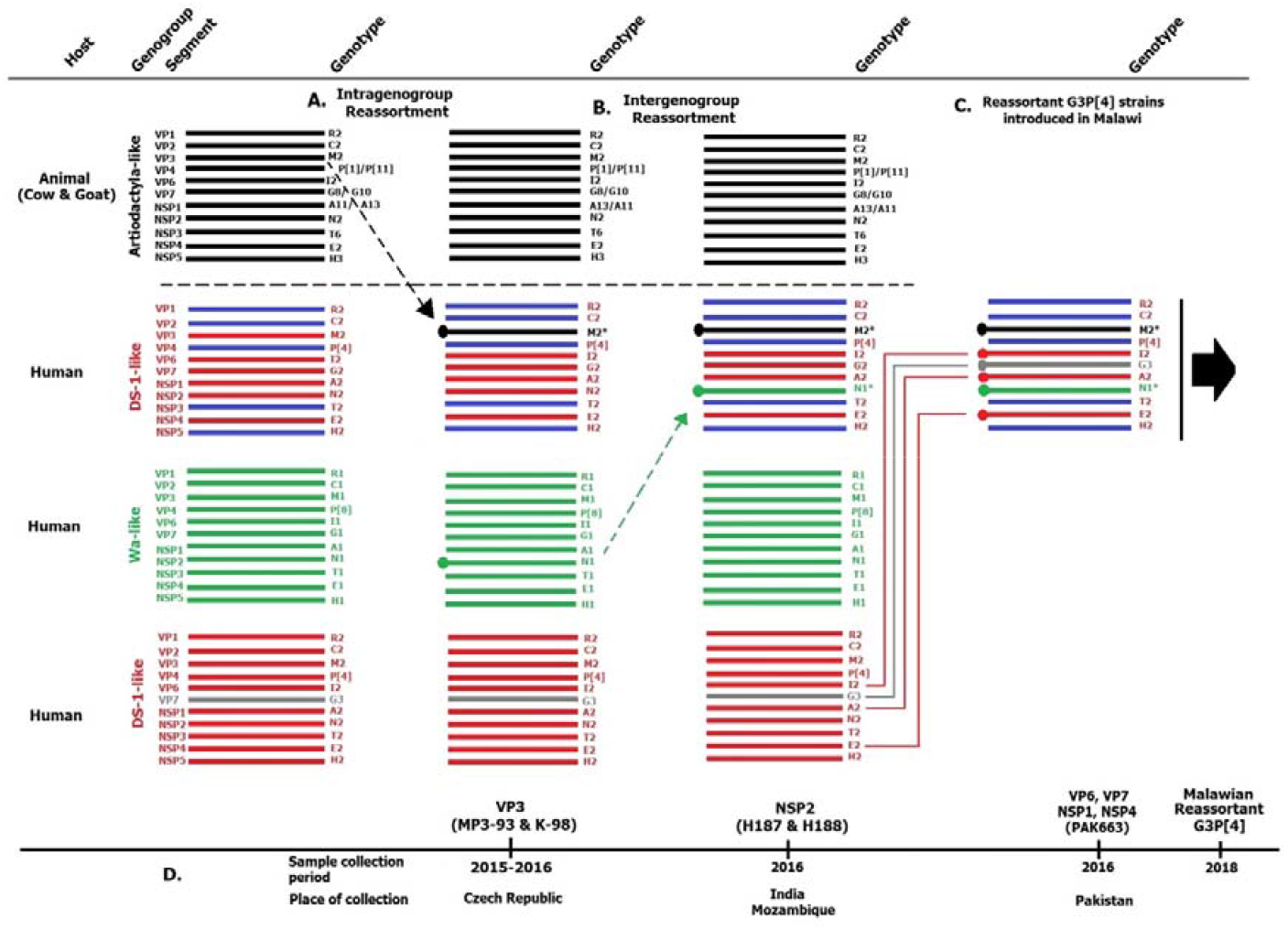
Reconstruction of the sequence of reassortment events that potentially lead to the emergence of the reassortant G3P[4] strains that were detected in Malawi. The reassortment events were hypothesised based on the genetic relationships between the genome segments of the reassortant DS-1-like G3P[4] rotaviruses that were detected in Malawi and those available in the NCBI Genbank to date. (A) The reassortant DS-1-like G3P[4] rotavirus strains that were detected in Malawi likely acquired their VP1, VP2, VP4, NSP1 and NSP5 genome segments from human DS-1-like rotaviruses and their VP3 from artiodactyl rotaviruses that circulated in countries where circulating rotaviruses have not been sequenced yet, or if done, their full-length genome sequences have not yet been deposited to the NCBI Genbank. (B) Both inter genogroup and intragenogroup reassortment events between Wa and DS-1-like rotaviruses were likely taking place from these unsampled regions, of which various progeny rotavirus populations were possibly generated. Reassortant DS-1-like strains that acquired their NSP2 through intergenogroup reassortment from Wa-like strains, which shared closest nucleotide similarity with rotaviruses that circulated in countries like Czech Republic between 2015 and 2016. (C) Various evolutionary events likely took place between the reassortant DS-1-like rotaviruses, DS-1-like G3P[4] and Wa-like G3P[8] rotaviruses that circulated in regions like Pakistan between 2014 – 2016, and in other unsampled regions that potentially led to various progeny rotavirus populations. One of the resultant progeny populations could be the reassortant DS-1-like G3P[4] strains from (B) that acquired NSP1, NSP4, VP6 and VP7 from the DS-1-like G3P[4] strains that were circulating in regions like Pakistan through complex reassortment events. The resultant reassortant DS-1-like G3P[4] rotaviruses were the ones that were exported to Malawi and caused diarrhoea disease in children that were hospitalised at Queen Elizabeth Central Hospital in Blantyre, Malawi from 2018, or circulated in unsampled regions. (D) The period and country where the closest related strains to the genome segments of the reassortant DS-1-like G3P[4] were detected. The horizontal line represents the 11 rotavirus genome segments. The colours represent the following: Green - Wa-like genome segments; Red - DS-1-like genome segments; Black - Artiodactyla-like genome segments; Grey - G3 genotype; Blue - Distinct genome segments.

### Amino acid substitutions within antigenic regions of G3P[8] have potential to drive vaccine escape variants

Antigenic regions of the rotavirus outer capsid proteins are critical for inducing neutralising antibodies (Estes et al. 2007). We assessed if there were amino acid differences between antigenic regions of the Malawian G3s strains to that of the G1P[8] Rotarix vaccine strain, and determined the structural conformational differences resulting from these mismatches. Due to the known amino acid mismatches between heterotypic rotavirus genotypes (Matthijnssens, Ciarlet, Heiman, et al. 2008), this analysis was limited to homotypic genotypes hence only the VP4 proteins of the G3P[8] strains were compared to that of the G1P[8] Rotarix vaccine strain (P[8] genotype). At least 32 out of the 36 amino acids that spans across the antigenic regions along the VP5* and VP8* subunits of the VP4 for the G3P[8] strains matched those of the Rotarix strain (Supplementary Table S3). The VP5* of the study G3P[8] strains (lineage III) were identical to that of Rotarix strain (lineage I) across all antigenic regions (Supplementary Table S3). However, differences were observed within the VP8*-1 and VP8*-3 antigenic regions. The E150D was the only substitution along the VP8*-1 antigenic region, whereas S125N, S131R and N135D substitutions occurred within the VP8*-3 antigenic region (Supplementary Table S4). The S131R amino acid substitution within the VP8*-3 resulted in a change from a charged amino acid with a potential of five hydrogen bonds to a polar amino acid with four hydrogen bonds. When we aligned the VP8* protein structures of the G3P[8] study strains against that of Rotarix strain, the superimposed structures revealed structural conformation differences within antigenic epitope region 8*-1 and 8*-3 specifically at positions 150 as well as 131, respectively (Supplementary Figure S4). The differences in the structural conformation of the antigenic region has potential to impact the neutralising ability of the Rotarix vaccine-induced antibodies against G3P[8] strains.

## Discussion

In this study, we have shown that G3 rotaviruses that re-emerged in 2017 in Malawi, almost twenty years after their last detection, became the predominant strains post-Rotarix vaccine introduction after replacing G1 and G2 rotaviruses as typical human strains. This finding is unique and in contrast with observations seen in most countries, including Australia, Italy, Hungary, Spain, Japan and Kenya where the emergence of G3 strains has mostly been due to equine-like G3 strains (Komoto et al. 2018; Dóró et al. 2016; Cowley et al. 2016; Luchs et al. 2019; Esposito et al. 2019; Mwanga et al. 2020). Through whole-genome sequencing, we showed that the re-emergence of G3 strains in Malawi was not associated with a single genotype, but rather four genotype variants; G3P[4] and G3P[6] with a DS-1-like constellation, G3P[8] with a Wa-like constellation and G3P[4] genotypes with a DS-1-like backbone containing a reassortant Wa-like NSP2 genome segment. Chronologically, these G3 rotaviruses circulated in three phases in Malawi whereby the DS-1-like G3P[4] strains were the first to emerge in November 2017 until August 2018 followed by a sporadic detection of DS-1-like G3P[6] in September 2018, Wa-like G3P[8] from December 2018 to August 2019 and reassortant DS-1-like G3P[4] from December 2018 until August 2019 with the Wa-like G3P[8] strains predominating in the latter period (2018 to 2019). Phylogenetic analysis revealed that multiple G3 lineages circulated in Malawi and that the re-emerging G3 strains did not arise due to stepwise evolution of the previously circulating strains, but rather through genomic reassortment and importation of strains from other countries. Altogether, these findings highlight the role of genome reassortment in driving rotavirus evolution and human mobility in disseminating rotavirus strains internationally.

Like other segmented viruses, such as influenza (Webster et al. 1992), rotaviruses frequently reassort their genomic segments, which increases their genetic diversity (Martínez-Laso et al. 2009). Although the majority of the human-associated G3 strains have a P[8] and Wa-like genotype constellation (Matthijnssens and Van Ranst 2012), most of the recently emerged G3 rotaviruses possess equine-like as well as DS-1-like rotavirus genome segments (Cowley et al. 2016; Dóró et al. 2016; Luchs et al. 2019; Komoto et al. 2018; Arana et al. 2016; Esposito et al. 2019). Whole genome sequencing of G3 rotavirus strains that were detected from other countries has shown that intergenogroup reassortment events between human and equine rotaviruses drove their emergence (Malasao et al. 2015). In contrast, the G3 strains that re-emerged in Malawi had typical human Wa-like and DS-1-like genetic constellations. These findings were similar to previous analysis of rotavirus genotypes that circulated in Malawi for over two decades from 1997 identified a diverse population of rotaviruses (at least 24 G and P genotype combinations) of which WGS revealed the majority had either a Wa-like or DS-1-like genetic backbone (Mhango et al. 2020; Jere et al. 2018). The four G3 variants that re-emerged in Malawi from 2017 also had either a Wa-like or DS-1-like genetic backbone but phylogenetic analysis revealed up to four lineages in each genome segment (one in all Wa-like and up to three in DS-1-like). This data demonstrates the wide diversity of G3 strains that co-circulated in Malawi within the two-year study period.

Further phylogenetic analyses of the re-emergent G3P[4] strains revealed that two populations circulated chronologically in Malawi. The DS-1-like G3P[4] strains that emerged first were genetically closest to sequenced G3 strains from Asia (Pakistan), whereas the reassortant DS-1-like G3P[4] strains, which emerged afterward were genetically distantly related to the former DS-1-like G3P[4] strains in seven genome segments. Our phylogenetic analysis suggested that the reassortant strains did not emerge from the DS-1-like G3P[4] that were first detected in Malawi between 2017 and 2018. In addition, the two G3P[4] populations did not share a recent common ancestor in NSP3, VP1, VP2 and VP4; thus, the reassortant DS-1-like strains were most likely not progenies of the DS-1-like G3P[4] strains that re-emerged first in Malawi and may have acquired these genome segments from elsewhere or could have been circulating independently at very low frequencies in such a way that could not be picked by our surveillance system. Indeed, three genome segments (NSP5, VP1 and VP4) of the reassortant DS-1-like G3P[4] strains clustered closely with strains from Europe (Czech Republic) instead of other contemporary DS-1-like G3 strains detected in Asia (Pakistan) or DS-1-like G3P[4] and non-G3 strains that circulated previously in Malawi. These findings suggested that these reassortant DS-1-like G3P[4] strains were potentially imported from other countries, although we cannot discount that these strains circulated previously in Malawi as we did not have sufficient sequenced G3 strains before their disappearance in the 1990s. Similarly, phylogenetic analysis of our reassortant DS-1-like G3P[4] showed that their Wa-like NSP2 (N1) genome segments were genetically similar to those from Czech Republic than those from Malawi while the VP3 genome segments showed a high nucleotide sequence similarity and clustered closely to Artiodactyl rotavirus strains characterised elsewhere. Considering our genomic surveillance could not pick up any of these animal-like genome segments, our findings suggested that the VP3 genome segments were not acquired in Malawi; rather, the reassortant strains were potentially seeded into Malawi already having this segment. Therefore, as none of the genome segments of the reassortant DS-1-like G3P[4] resemble strains sequenced from Malawi at any time, it is likely that the reassortant strains containing a Wa-like NSP2 acquired their VP7, VP6, NSP1 and NSP4 genome segments from G3P[4] through intragenogroup reassortment elsewhere prior to their introduction into Malawi in 2018. Together, these findings suggest that the typical and reassortant G3 strains that re-emerged in Malawi were potentially imported, suggesting that importation of rotavirus strains may be an overlooked, but key driver for reseeding genotypes in different countries (Mhango et al. 2020; Jere et al. 2018).

Emerging virus strains have been associated with mutations that render vaccines and other therapies, such as monoclonal antibodies, less effective as seen with the severe acute respiratory syndrome coronavirus 2 (SARS-CoV-2) (Starr et al. 2021). Previous studies have shown that the VP5* and VP8* outer capsid proteins play a significant role in inducing neutralising antibodies against rotaviruses (Ludert et al. 2002; Park et al. 2021; Ruggeri and Greenberg 1991; Zhao et al. 2015). The VP5* protein of the Malawian G3P[8] strains were 100% conserved when compared to that of Rotarix vaccine, consistent with previous studies that have reported that the VP5* is a highly conserved protein (Rasebotsa et al. 2020; Mwangi et al. 2020). However, we identified E150D nonsynonymous substitutions within the VP8*-1 antigenic region and S125N, S131R and N135D substitutions within VP8*-3 antigenic region when we compared the VP4 (VP8*sub-unit) segments of our G3P[8] strains to that of the strain used in the Rotarix rotavirus vaccine. These substitutions have been associated with lineage III strains that are currently predominant in eastern and southern African countries where the Rotarix vaccine is used (Rasebotsa et al. 2020; Mwangi et al. 2020; Maringa et al. 2021). We speculate that the structural changes we observed in the VP8*-1 and VP8*-3 antigenic regions could reduce the binding of the vaccine-induced antibodies thereby reducing the vaccine effectiveness. Further studies are required to investigate the impact of these nonsynonymous amino acid changes to generate a complete map of vaccine escape mutants and how they affect antibody neutralisation.

Our study has some limitations. Due to the destruction of the historical stool sample collected through our rotavirus surveillance system as part of the polio containment campaign in Malawi, we included only a few G3P[8] strains from the pre-rotavirus vaccine period sequenced earlier as we were unable to sequence additional pre-vaccine strains. Therefore, although we have shown that the re-emerged G3P[8] strains in Malawi are unlikely to have emerged from those circulating in Malawi in the pre-vaccination era, we cannot completely rule out that these strains did not emerge from other unsampled strains circulating during this period. We also understand that the contextual rotavirus sequences obtained from GenBank are sparse as sequencing of rotavirus strains is not routinely performed in many countries, which leads to massive surveillance gaps globally. Because of this, we could only infer that the re-emerged G3 strains in Malawi were imported but we cannot say with absolute certainty the country of origin for these strains. In addition, we halted our rotavirus surveillance from 2020 to 2021 due to the COVID-19 pandemic, which prevented us from investigating G3 rotaviruses over a longer time period after their re-emergence. Regardless, we were able to generate representative G3 strains across the two years of rotavirus surveillance that covered the period when G3 genotypes were detected at high frequency.

To our knowledge, our study provides the first comprehensive and systematic genomic characterization of the re-emerging G3 rotavirus from Africa. Our findings demonstrates that four G3 rotaviruses genotype variants resembling typical human rotaviruses re-emerged after Rotarix rotavirus vaccine introduction twenty years after disappearing for almost twenty years in Malawi, and their emergence appear to be likely driven by importation from other countries. Our findings highlight the role of human mobility in driving the dissemination and temporal dynamics of circulating rotaviruses internationally and demonstrates the importance of robust rotavirus surveillance and whole genome sequencing to monitor strain dynamics to inform infection prevention and control strategies in high disease burden settings.

## Methods

### Ethical approval

Informed consent was obtained from all the mothers or legal guardians for the children who were involved in this study. This study was conducted according to the guidelines of the Declaration of Helsinki and approved by the Research Ethics Committee of the University of Liverpool, Liverpool, UK (000490) and the National Health Sciences Research Committee, Lilongwe, Malawi (#867).

### Sample collection, rotavirus genotyping and selection of rotavirus strains

Stool samples were collected from children <5 years old who presented with acute gastroenteritis to the Queen Elizabeth Central Hospital (QECH) in Blantyre, Malawi through a rotavirus surveillance platform which has been on-going since 1997 (Mhango et al. 2020). Acute gastroenteritis was defined as the passage of at least 3 loose, or looser-than-normal stools every 24 hours for less than one week. Presence of rotaviruses in stool samples was confirmed using Rotaclone^®^ Enzyme Immunosorbent Assay (Rotaclone^®^, Meridian Bioscience, Cincinnati, OH, USA). The VP7 and VP4 genotypes for rotavirus positive samples were assigned using a multiplex heminested reverse transcriptase polymerase chain reaction (RT-PCR) as described previously (Mhango et al. 2020; Iturriza-Gomara et al. 1999). At least one specimen containing rotavirus of G3 genotype was selected each calendar month from November 2017 to August 2019 for sequencing (*n=27*) (Supplementary Figure 1). Whole genomes for three G3 strains that circulated between 1997 to 1999 collected and sequenced from our previous studies were also analysed (Jere et al. 2018; Cunliffe et al. 2010).

### Extraction of rotavirus double-stranded RNA and synthesis of complementary DNA

Rotavirus dsRNA was extracted and purified as previously described (Jere et al. 2011, 2018). To remove contaminating DNA, the extracted dsRNA was precipitated with lithium chloride (Sigma-Aldrich, Dorset, UK) for 16 hrs at 4°C and treated with DNase I (Sigma-Aldrich, Dorset, UK) as previously described (Jere et al. 2018). Purified dsRNA was quantified on Qubit 3.0 fluorometer (Life Technologies, CA, USA). A 1% 0.5 X Tris borate ethylenediaminetetraacetic acid (TBE) agarose gel (Sigma-Aldrich, Dorset, UK) stained with SYBR green (Sigma-Aldrich, Dorset, UK) electrophoresis was used to check the integrity of the extracted dsRNA and was visualised on a BioDoc transilluminator. Complementary DNA (cDNA) was synthesised using the Maxima H Minus Double-Stranded cDNA Synthesis Kit (Thermo Fisher Scientific, Waltham, MA) and purification was done using the MSB^®^ Spin PCRapace (Stratec) Purification Kit as previously described (Mwangi et al. 2020; Rasebotsa et al. 2020).

### DNA library preparation and whole genome sequencing

The Nextera XT DNA Library Preparation Kit (Illumina, San Diego, CA, USA) was used to prepare DNA libraries following the manufacturer’s instructions. Briefly, the Nextera^®^ transposome enzyme was used to target genomic DNA which was amplified using a limited cycle PCR. AMPure XP magnetic beads (Beckman Coulter, Pasadena, CA, USA) and 80% alcohol were used to clean-up the DNA libraries. Qubit 3.0 fluorometer (Invitrogen, Carlsbad, CA, USA) was used to quantify the cleaned-up DNA libraries. The fragment size and quality of libraries were assessed on Agilent 2100 BioAnalyzer^®^ (Agilent Technologies, Waldbronn, Germany). Paired end nucleotide sequences were then generated on a MiSeq^®^ sequencer (Illumina, San Diego, CA, USA) at the University of the Free State-Next Generation Sequencing (UFS-NGS) Unit, Bloemfontein, South Africa as previously described (Mwangi et al. 2020; Rasebotsa et al. 2020).

### Sequence assembly and whole genome genotype determination

We checked the quality of the whole genome sequencing data using FASTQC (de Sena Brandine and Smith 2019) and selected samples with quality score >30 for subsequent analysis. Illumina adapter sequences were trimmed from the raw FASTQ sequence data using BBDuk trimmer (version 2) (https://sourceforge.net/projects/bbmap/) embedded in Geneious Prime software (version 2020.1.1) (Kearse and Sturrock, n.d.). Consensus sequences were generated through mapping of trimmed Illumina reads to prototype rotavirus Wa-like (accession numbers JX406747.1 - JX406757.1) and DS-1-like (accession numbers HQ650116.1 - HQ650126.1) genogroup reference strains by Geneious Read Mapper (version 6.0.3) with the medium sensitivity and iteratively fine-tuning parameters five times in Geneious Prime software (Kearse and Sturrock, n.d.). The Geneious consensus tool was used to call the total quality consensus by selecting a 60□% highest quality threshold. The gene annotation and prediction tool in Geneious Prime was used to annotate regions of low coverage (<200). To validate the consensus sequences generated by mapping reads to reference sequences, we generated *de novo* assemblies using Iterative Virus Assembler (IVA, version 1.0.3) pipeline (Hunt et al. 2015) for comparison. We assigned the genotypes of each assembled genome segment using the Virus Resource Pathogen (ViPR) online server for viral genotyping (Pickett et al. 2012).

### Phylogenetic analysis

To compare our study strains to G3 rotaviruses characterised globally, we obtained the VP7 genome segment of G3 strains from the Virus Variation Resource in the GenBank (Hatcher et al. 2017). We selected genomic segments with a complete open reading frame (ORF) and aligned them using MUSCLE (version 3.8.1551) (Edgar 2004). Maximum likelihood phylogenetic trees were then generated in MEGAX (version 10.1.8) with generalised time reversible (GTR) and Gamma heterogeneity DNA models. We performed 1000 bootstraps to assess the reliability of the branching order and partitions in the phylogeny. Annotation of the phylogenetic trees was done using Microreact online server (Argimón et al. 2016). To define lineages for VP7, VP4 and DS-1-like genome segments (genotype 2), we used well-known lineage definition frameworks based on Sadiq et al 2019, Rasebotsa et al 2020 and Agbemabiese et al. 2019 work, respectively. Representative reference nucleotide sequences for DS-1-like genotypes for each genome segment were obtained from the Virus Variation Resource in the GenBank database (Hatcher et al. 2017). As there is no well-known lineage definition framework for Wa-like genome segments (genotype 1), global as well as local genotype 1 sequences sampled across the pre-vaccine and post-vaccine periods in Malawi were used as references. The nucleotide sequences for the ORFs of our study strains and reference strains were multiple aligned using MUSCLE (version 3.8.1551) (Edgar 2004). Once aligned, the DNA test models in MEGAX (version 10.1.8) (Tamura, Stecher, and Kumar 2021) were used to identify the optimal evolutionary model that best fit the data for each segment. According to the corrected Akaike Information Criterion (AIC’c) as previously described (Kumar et al. 2018), the GTR model with Gamma heterogeneity across nucleotide sites was selected and 1000 bootstraps were used to assess the reliability of the branching order and partitioning during the construction of maximum likelihood trees (Felsenstein 1988).

### Inference of the time to the most recent common ancestor

To estimate the most recent common ancestor (tMRCA) for each genome segment, we utilised nucleotide sequences of all Wa-like and DS-1-like strains that circulated between 1997 to 2019 in Malawi. We did not do genome-specific analysis for the G3 and P[6] genotype of the VP7 and VP4, respectively, because we did not have sufficient sequences to conduct the analysis. The reassortant Wa-like NSP2 (N1 genotype) genome segments for the double reassortant G3P[4] study strains were analysed together with other Wa-like NSP2 genome segments. In brief, we aligned genomic segments of previously circulating Wa-like and DS-1-like strains with the Wa-like and DS-1-like G3 segments, respectively, using MAFFT (version 7.487) (Katoh and Standley 2013). The alignments were trimmed at the 3’ and 5’ prime ends in order to generate sequences of equal lengths while preserving the integrity of the ORF as a pre analytical process. Trimmed alignments were then used to generate time-resolved trees using tree-time (version 0.8.0) (Sagulenko, Puller, and Neher 2018) and the trees were subsequently visualised and annotated using Auspice (version 2.23.0) (Hadfield et al. 2018). We exported the time-resolved trees from Auspice and visualised them using R (version 4.0.3). We used ladderized using the ‘ladderize’ function in ape (version 5.6.2) (Revell 2012) and rooted the tree based on an outgroup sequence using the ‘root’ function implemented in phytools (version 0.7.70) (Revell 2012). We estimated the phylogenetic root-to tip distance based on the sum of branch lengths (transformed to represent time in days) using ‘distRoot’ function in adephylo (version 1.1.11) (Jombart, Balloux, and Dray 2010) and visualised the tree annotated with the genotypes for each strain using the ‘plot.phylo’ function in the ape (version 5.6.2) package (Paradis and Schliep 2019).

### Structure comparison between the outer capsid VP4 proteins of the G3P[8] and Rotarix vaccine G1P[8] strains

To compare the antigenic sites of the VP4 of G3P[8] and that of Rotarix G1P[8] strains, we aligned their VP4 amino acid sequences using MAFFT (version 7.487) (Katoh and Standley 2013). We targeted the VP5* and VP8* antigenic regions and extracted antigenic sites from the alignments in MEGAX (version 10.1.8) (Tamura, Stecher, and Kumar 2021). To investigate the impact of amino acid substitutions within the antigenic sites on the structural conformation of the neutralising epitopes within the VP4 protein, we selected a representative VP4 amino acid sequences for G3P[8] strains and conducted protein modelling using Modeller (version 9.25) (Eswar et al. 2006). We selected three model structures with the highest Z-dope score and conducted a structural assessment using SWISS-MODEL server. The protein structures were visualised and annotated using PyMOL (version 2.4.1) (DeLano 2002).

## Data Availability

The clinical data presented in this study are available on request from the corresponding author. The data are not publicly available due to ethical restrictions. The whole-genome sequencing data generated for genome segments utilised in this project were submitted to the NCBI database under accession numbers ON791851-792171.

## Author Contributions

V.N.N., F.E.D, M.M.N., and K.C.J conceived, designed and sought funding for the study. M.I-G., N.A.C., O.K., and K.C.J. collected clinical data and stool samples. C.M., J.J.M., E.C., P.M. M.M.N., and K.C.J. performed the laboratory work. C.M., C.C., and K.C.J. carried out the statistical and bioinformatics analysis. C.C., A.W.K., and K.C.J. supervised the study. C.M., C.C., and K.C.J. drafted the manuscript. M.M.N generated the whole genome sequence data. C.M., A.B., E.C, J.J.M., O.K., C.M-B., K.G.B., B.K., K.C.J., C.M.D., M.D.E., A.D. S., P.M. M.I.-G, N.A.C., V.N.N., A.W.K., F.E.D., M.M.N, C.C., and K.C.J contributed to interpretation of the data and writing the manuscript. All authors have read and approved the final manuscript.

## Acknowledgments

We acknowledge the support of the laboratory staff at the Malawi-Liverpool-Wellcome Trust Clinical Research Programme, clinical research team and the study participants. We also acknowledge the University of Free State Next Generation Sequencing (UFS-NGS) unit staff for sequencing the G3 strains as part of the African Enterics Viral Genome Initiative (AEVGI).

