## Supplementary Figure S1 for "Comparative whole genome analysis reveals re-emergence of typical human Wa-like and DS-1-like G3 rotaviruses after Rotarix vaccine introduction in Malawi"

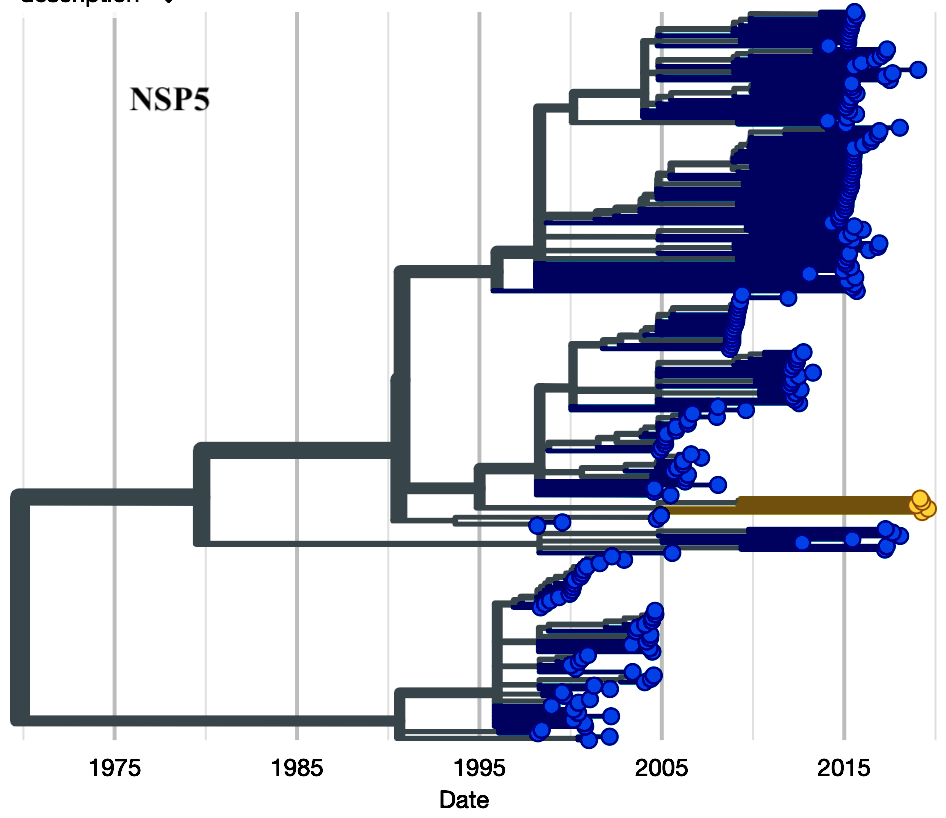

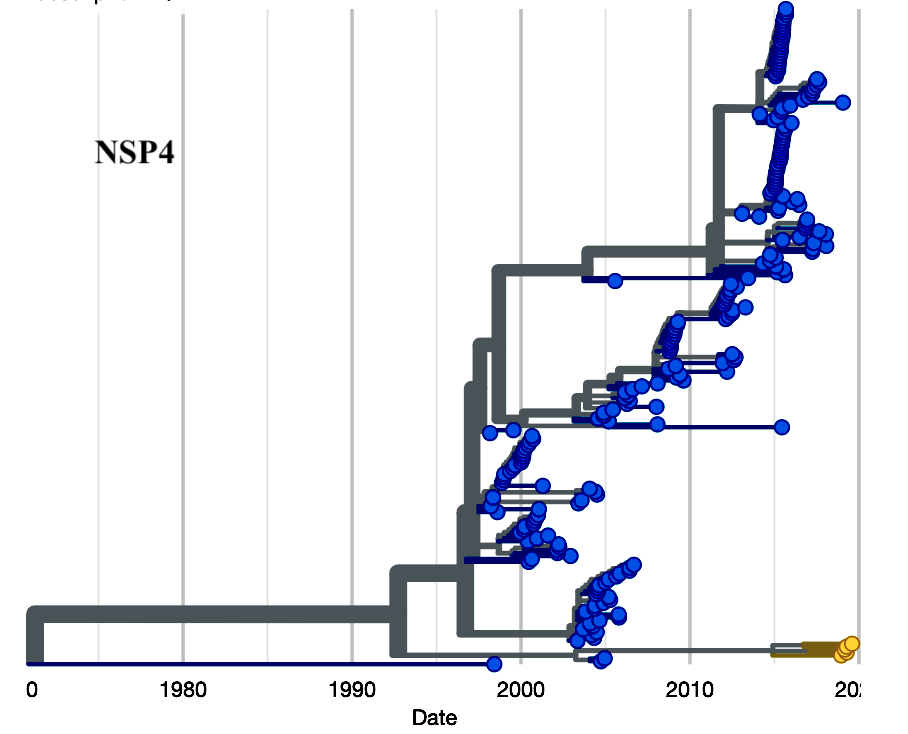

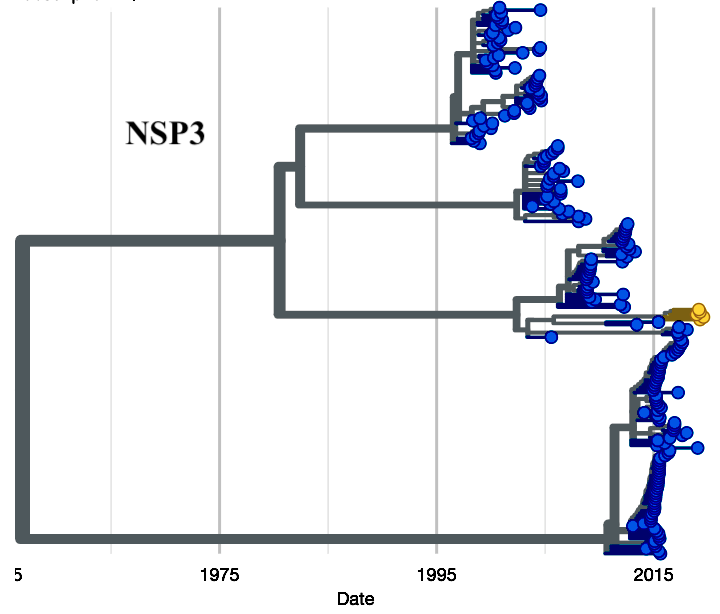

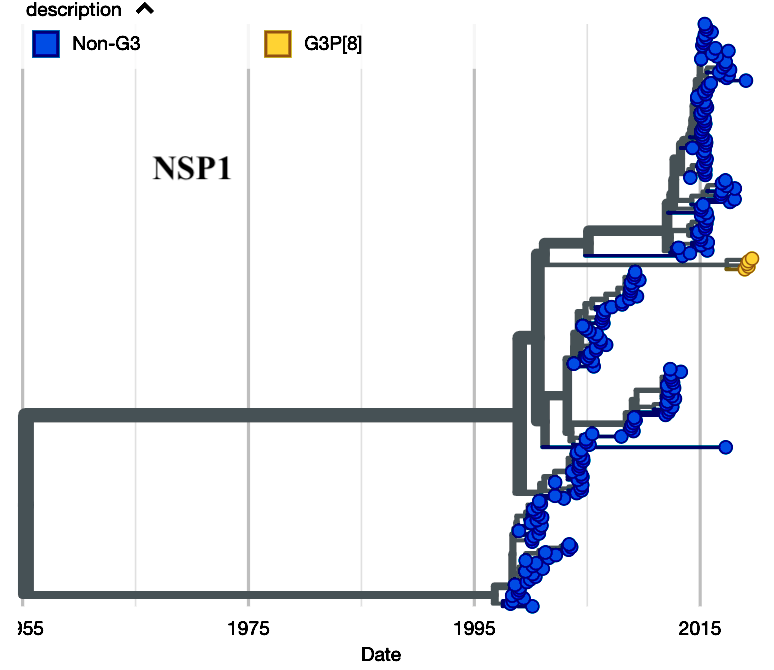

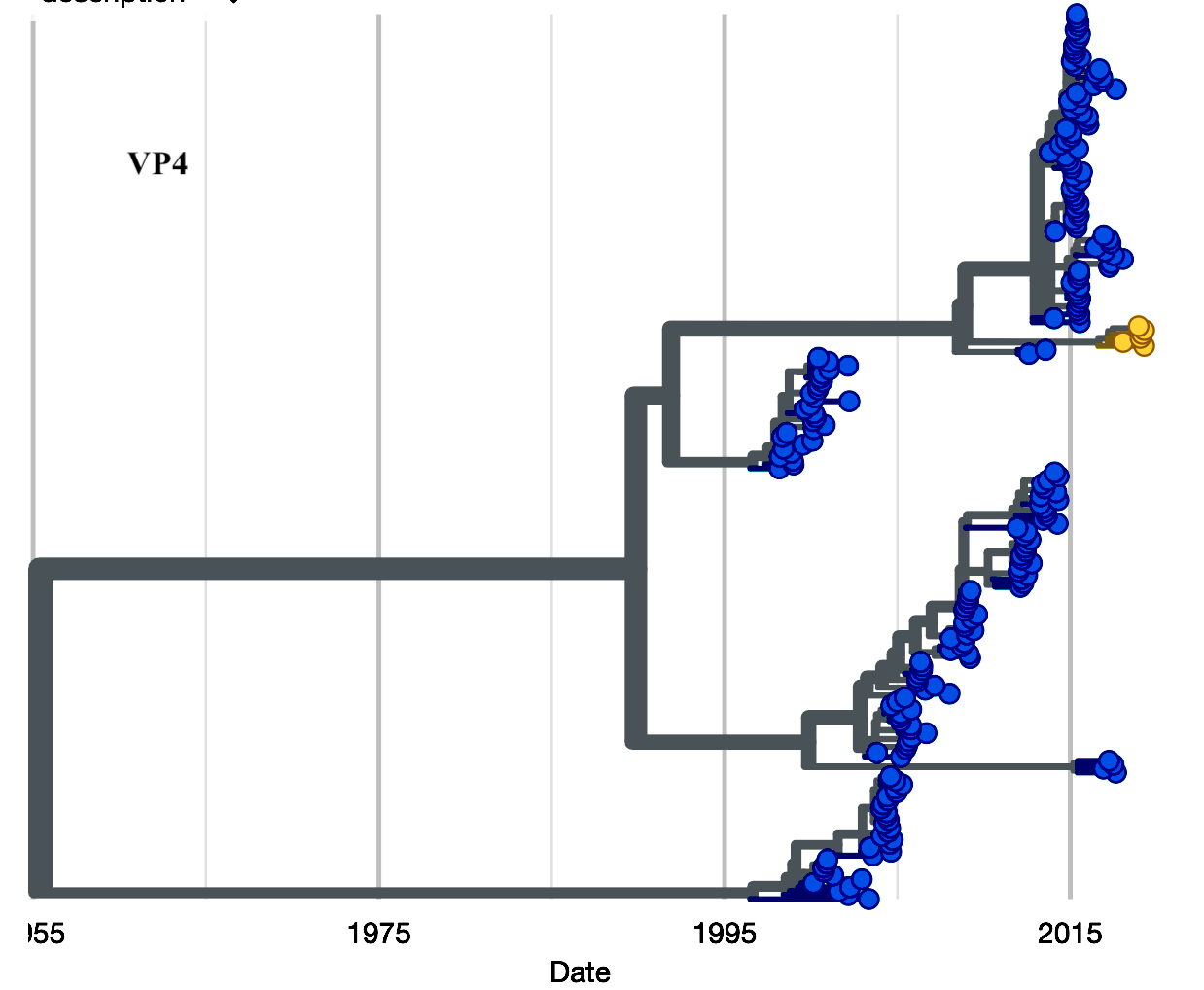

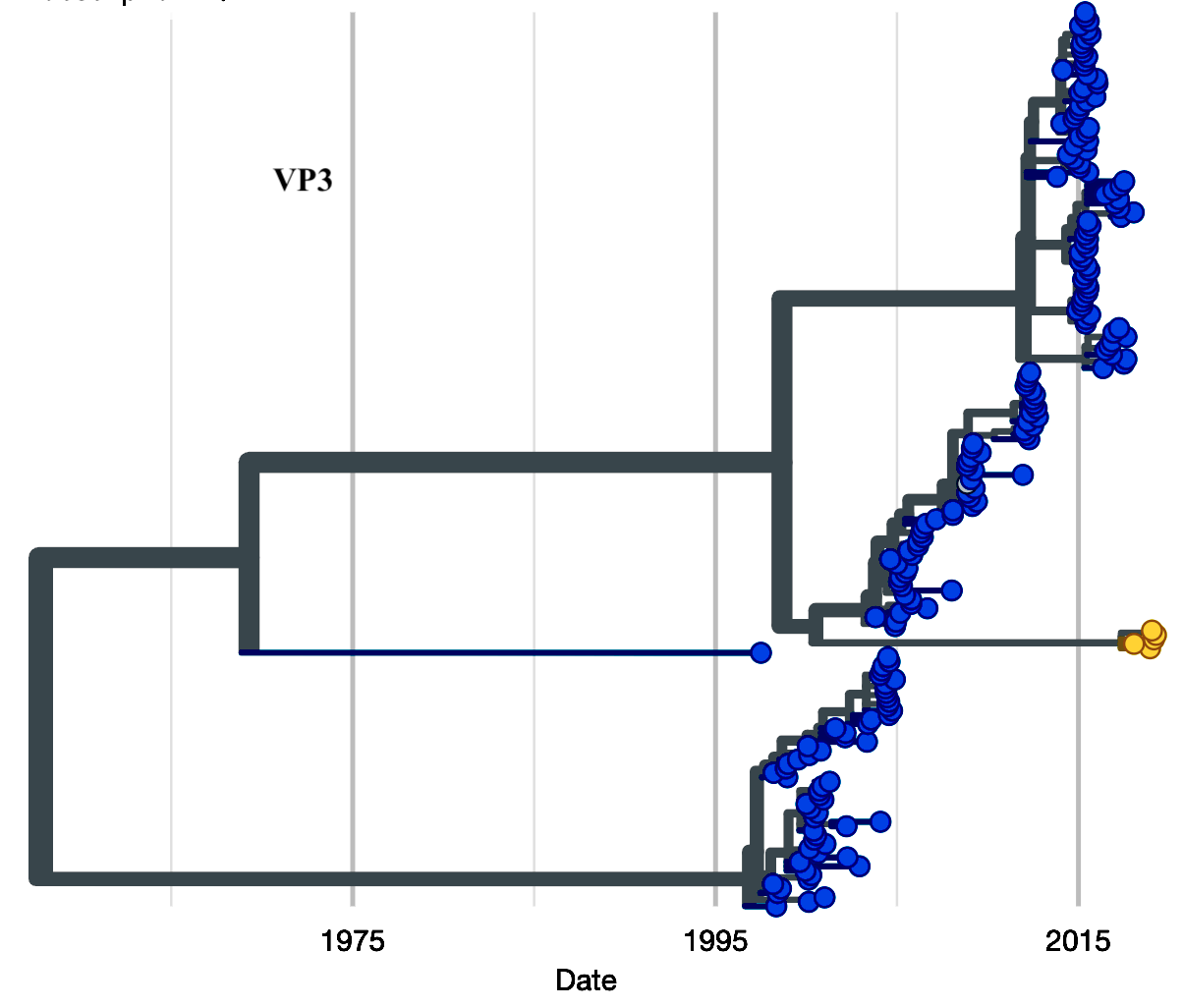

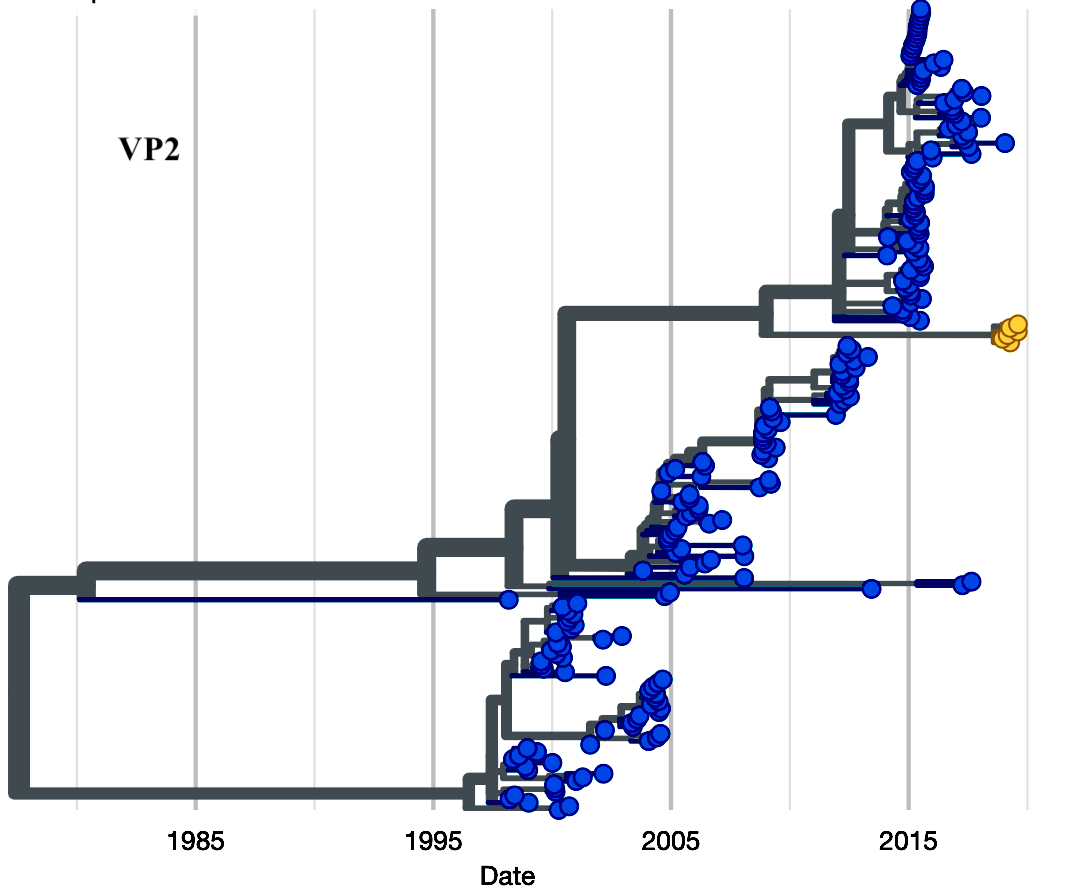

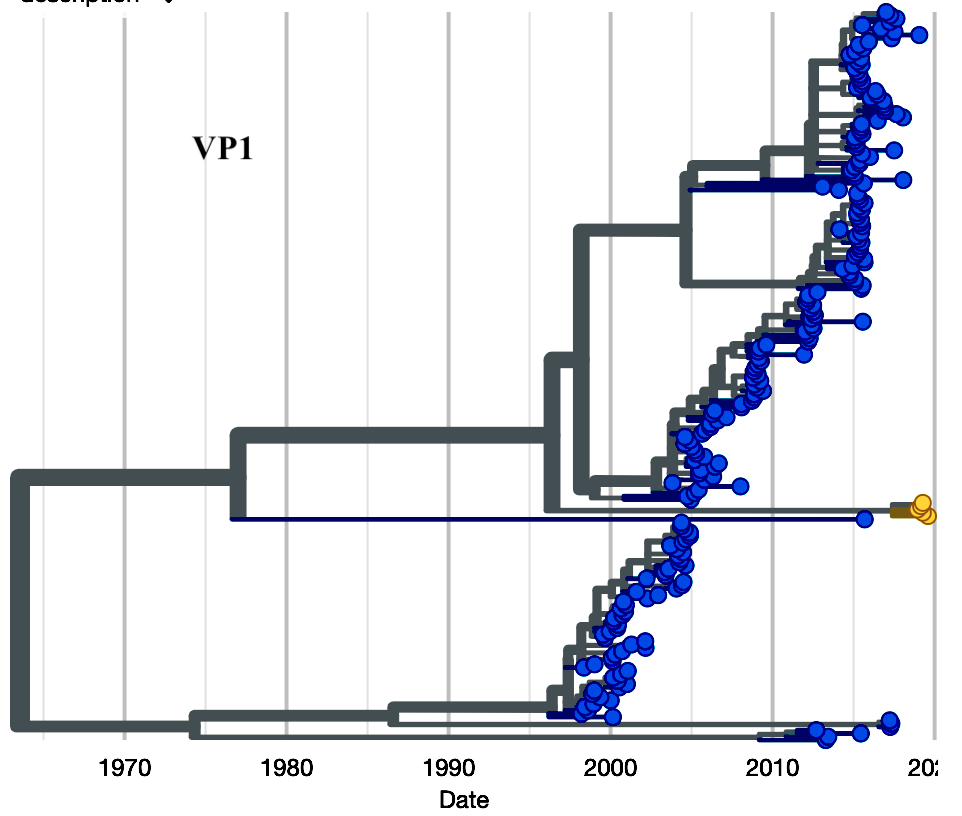

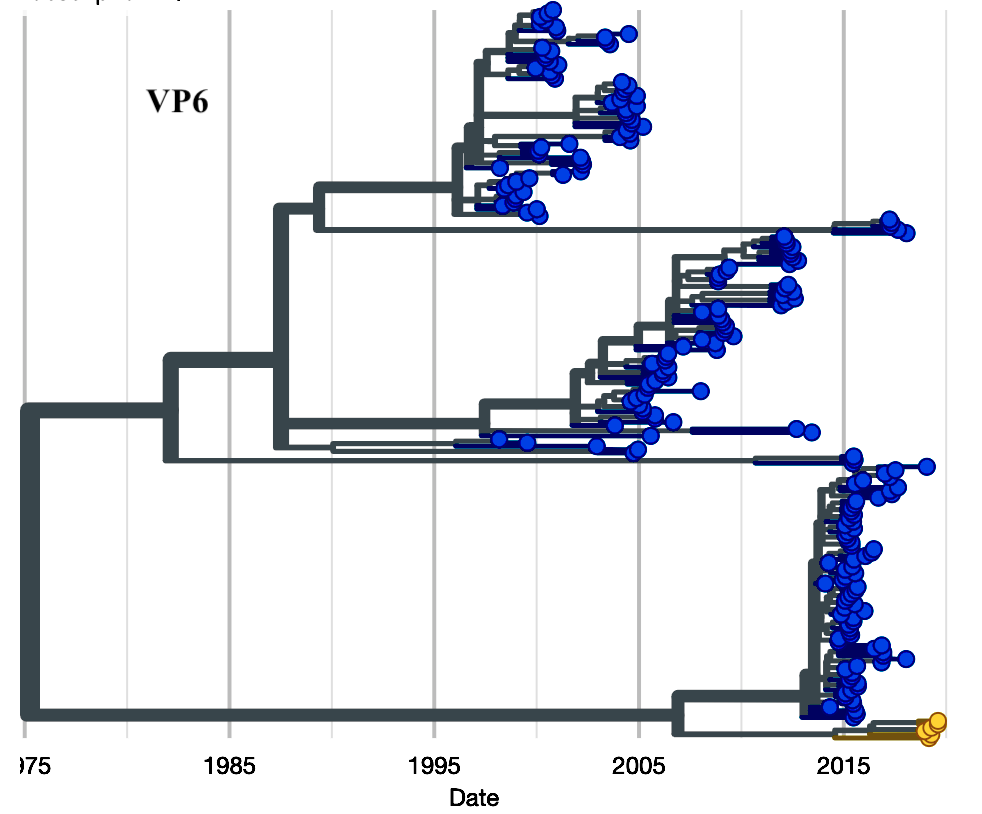

**Supplementary Figure S1.** **Time resolved phylogenetic trees generated using Nexstrain for all Wa-like genome segments detected in Malawi from 1997 to 2019 and Wa-like genome segments associated with re-emergent G3 strains**. The previously circulating Wa-like segments were used to estimate time to the most recent ancestor for the Wa-like G3 strains in relation to locally circulating Wa-like genome segments in Blantyre, Malawi. Genome segments associated with G3 rotavirus strains are annotated in yellow tips while non-G3 strains are annotated in blue tips. The VP4 segment only includes the P[8] genotype associated with G3 as well as other non-G3 strains characterized between 1997 and 2019. The rest of the genome segments are genotype 1 (Wa-like) only.
