## Supplementary Figure S2 for "Comparative whole genome analysis reveals re-emergence of typical human Wa-like and DS-1-like G3 rotaviruses after Rotarix vaccine introduction in Malawi"

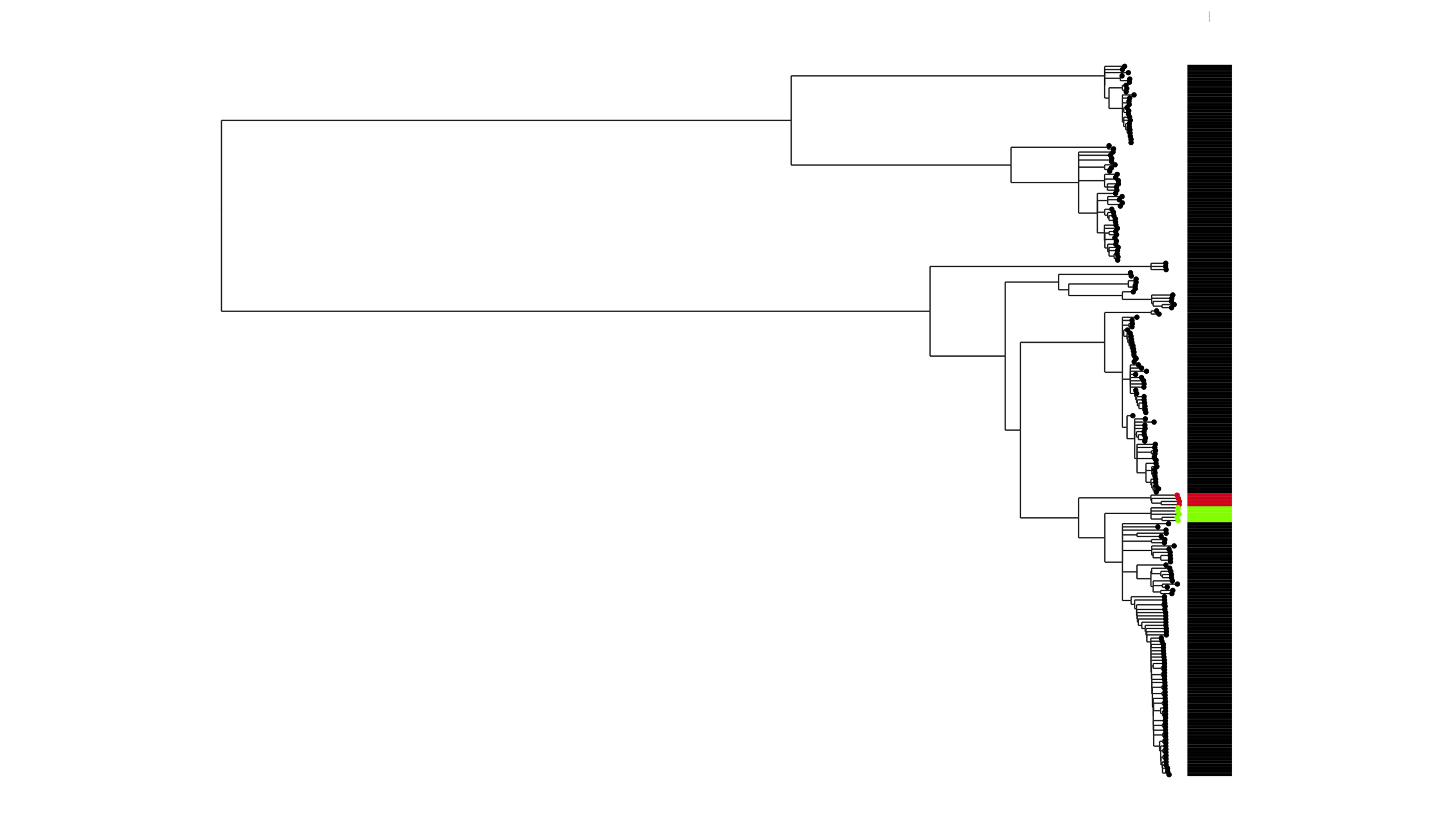

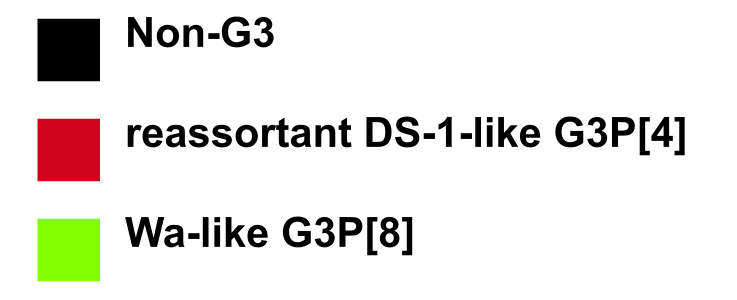

**Supplementary Figure S2**. **Maximum likelihood (ML) phylogenetic tree of all N1 NSP2 genotypes associated with G3 as well as non-G3 strains characterised in Malawi.** Only strains with a complete open reading frame were included in the analysis. The GTR evolutionary model with Gamma heterogeneity across nucleotide sites was used for phylogenetic inference. Bootstrap values ≥70% are shown adjacent to each branch node. The trees were out grouped at RVA/Pigeon-wt/JPN/P0-13/1989/G18P[17] but it was removed in the final tree for better visualization. Malawian Wa-like G3P[8] and reassortant DS-1-like G3P[4] strains are denoted by green and red colours respectively.
