## Supplementary Figure S3 for "Comparative whole genome analysis reveals re-emergence of typical human Wa-like and DS-1-like G3 rotaviruses after Rotarix vaccine introduction in Malawi"

a. **VP7**

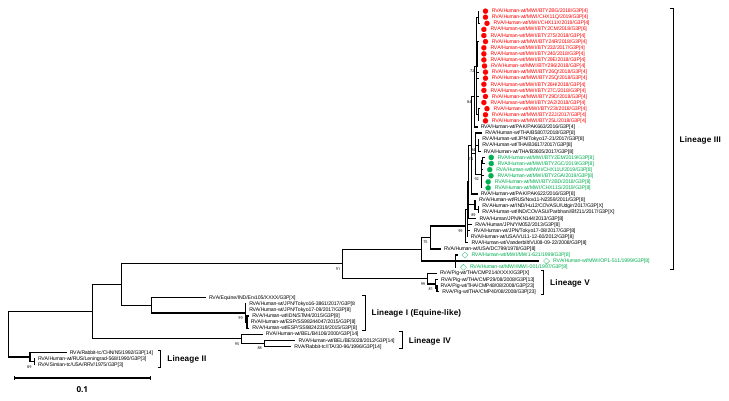

b. **VP6**

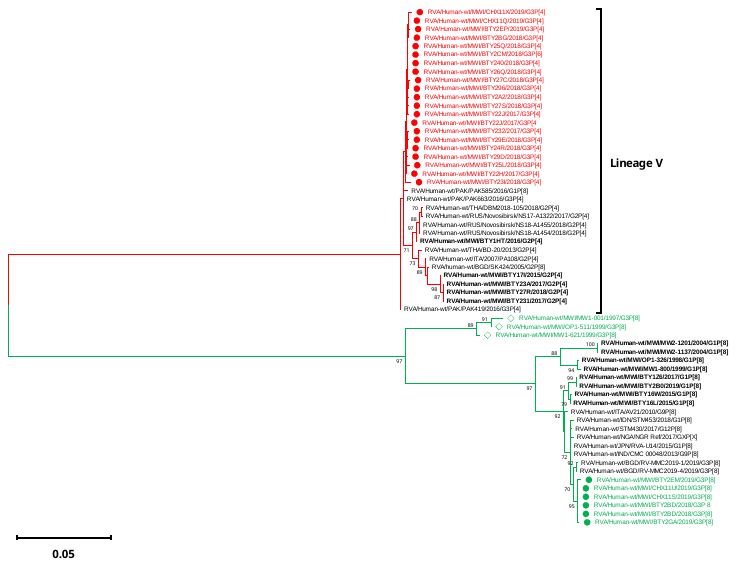

c. **VP4**

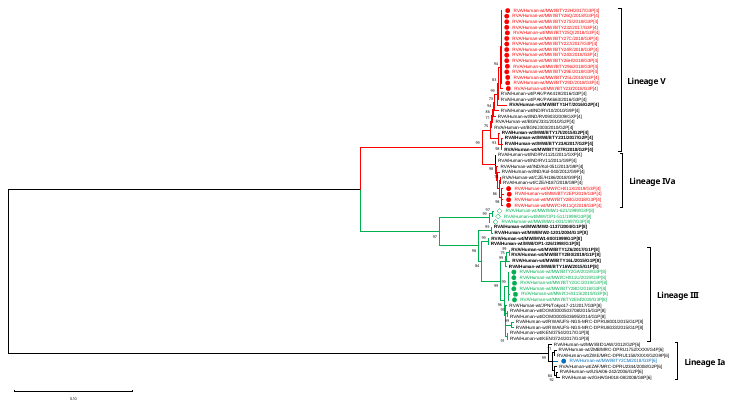

d. **VP2**

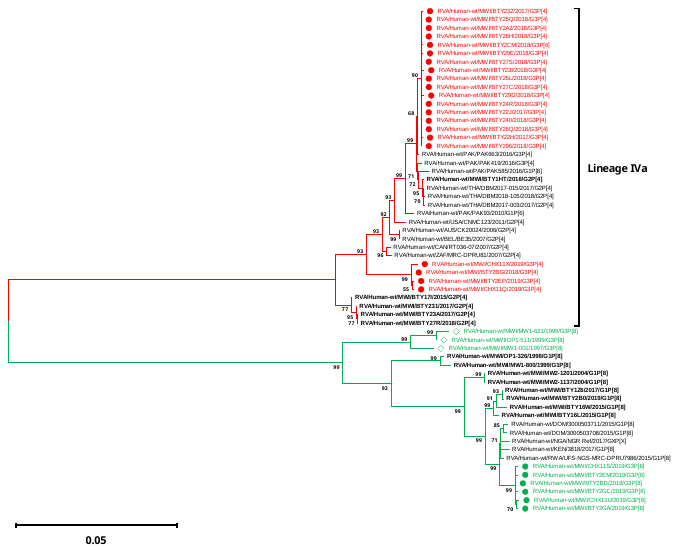

e. **VP1**

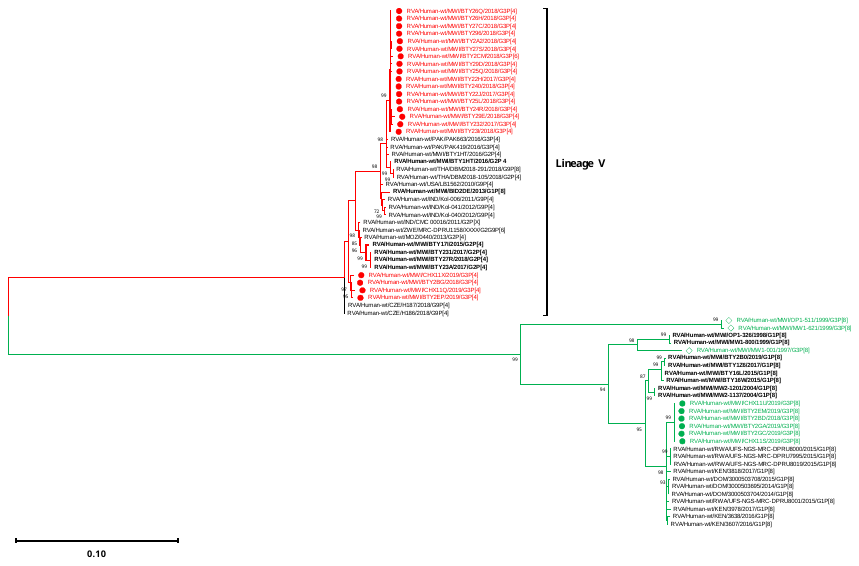

f. **NSP1**

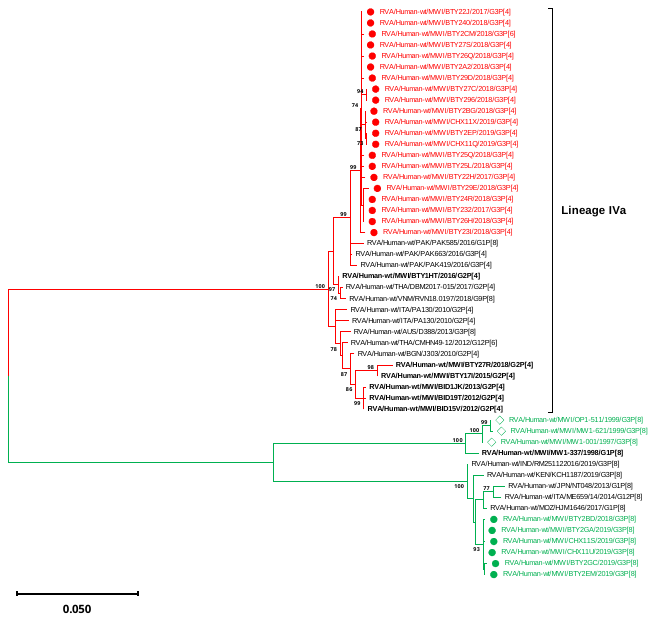

g. **NSP3**

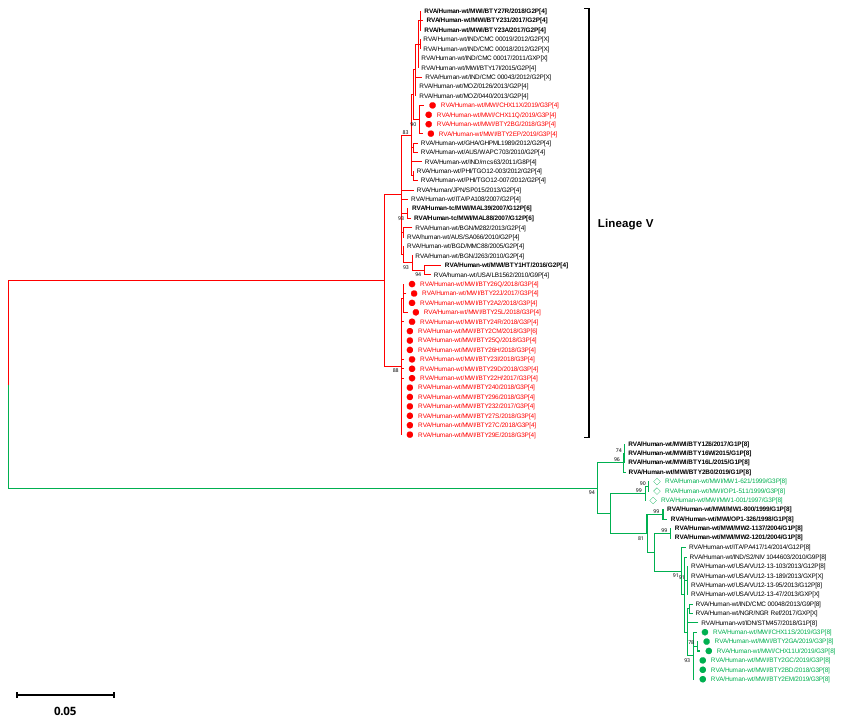

h. **NSP4**

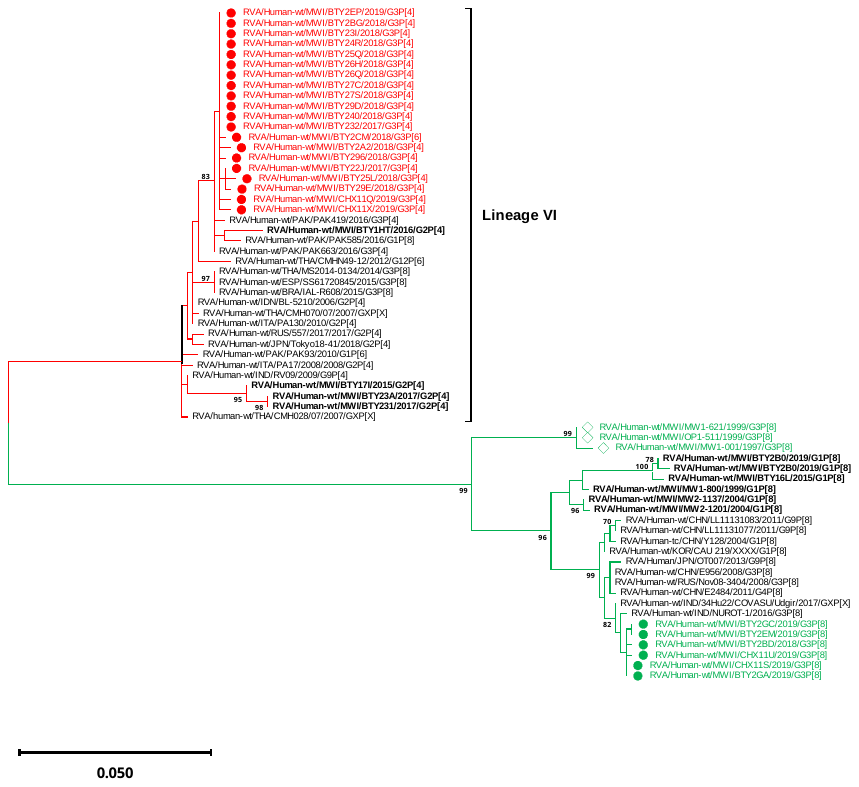

i. **NSP5**

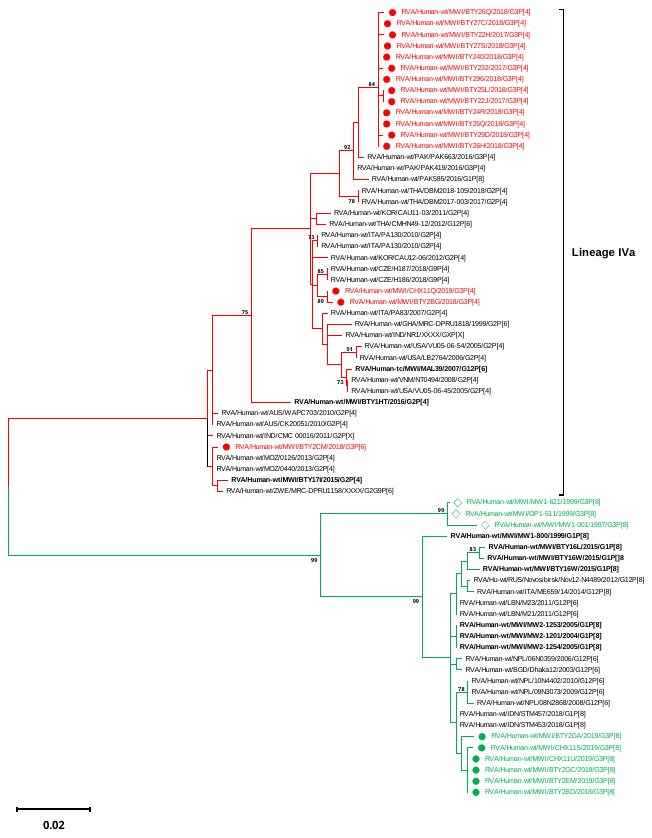

**Supplementary Figure S3**. **Maximum likelihood (ML) phylogenetic trees of Malawian G3 strains with global reference strains sharing a high nucleotide sequence similarity to Malawian strains**. Only strains with a complete open reading frame were included in the analysis. The GTR evolutionary model with Gamma heterogeneity across nucleotide sites was used for phylogenetic inference. Bootstrap values ≥70% are shown adjacent to each branch node. Malawian Wa-like and DS-1-like G3 strains are denoted by green and red colours respectively. Circles represent post-vaccine strains while diamonds represent pre-vaccine strains (A-H). Shows ML trees for VP7, VP4, VP6, VP1-VP2, NSP1, NSP3-NSP5 with a global lineage definition system for VP7, VP4 and DS-1-like genome segments while Wa-like genome segments have global as well as local reference strains
