## Supplementary Figure S4 for "Comparative whole genome analysis reveals re-emergence of typical human Wa-like and DS-1-like G3 rotaviruses after Rotarix vaccine introduction in Malawi"

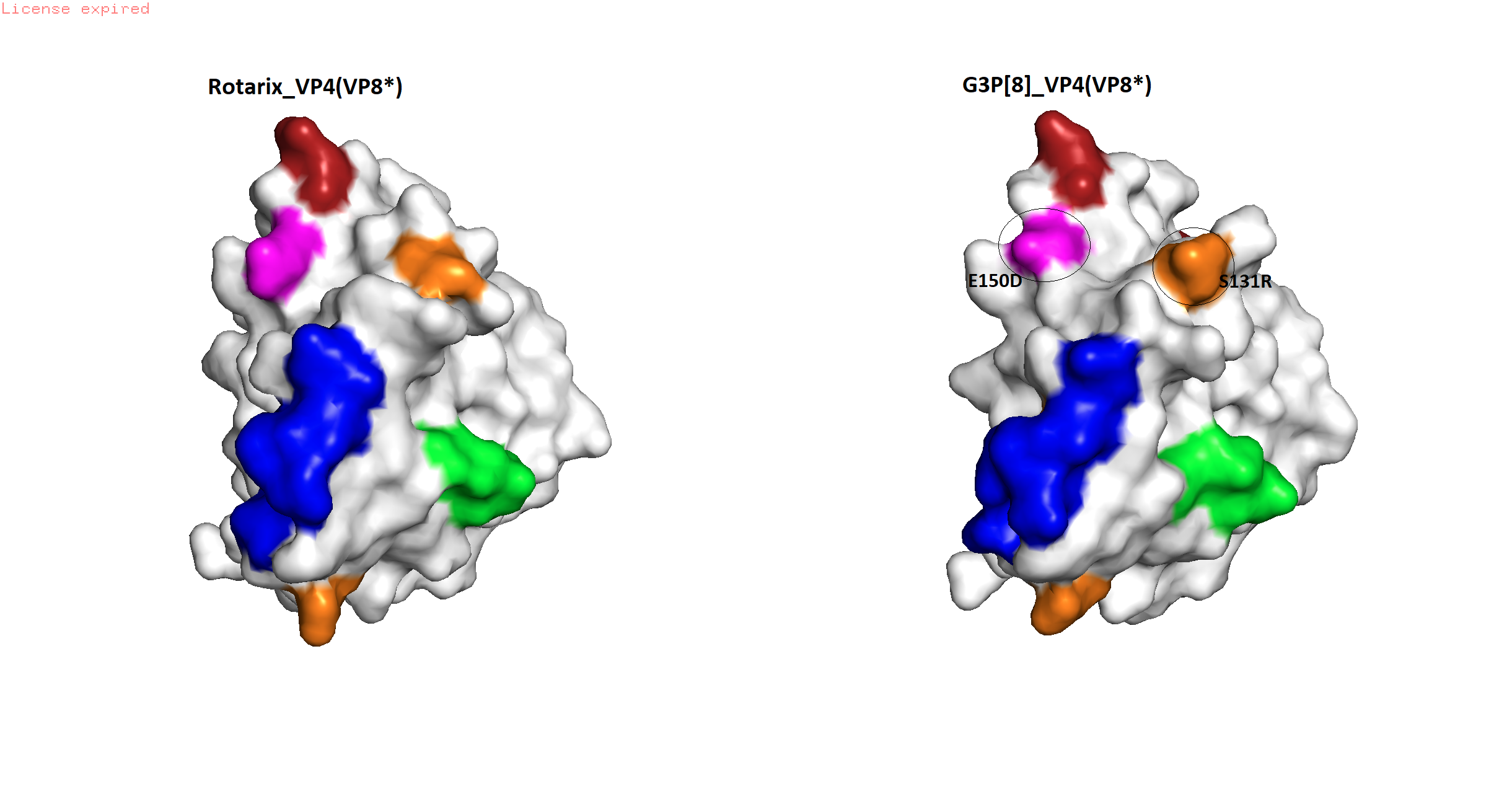

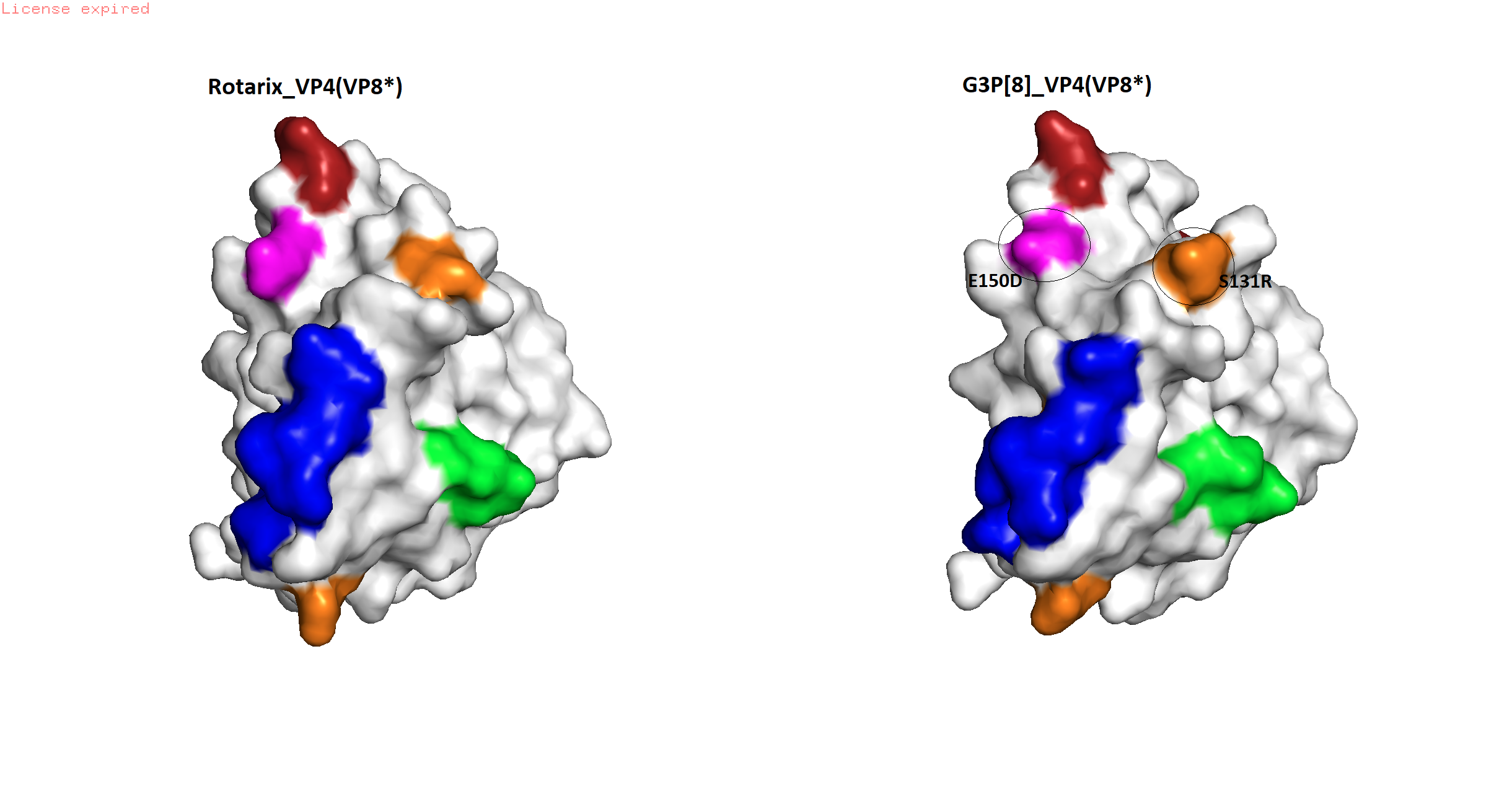

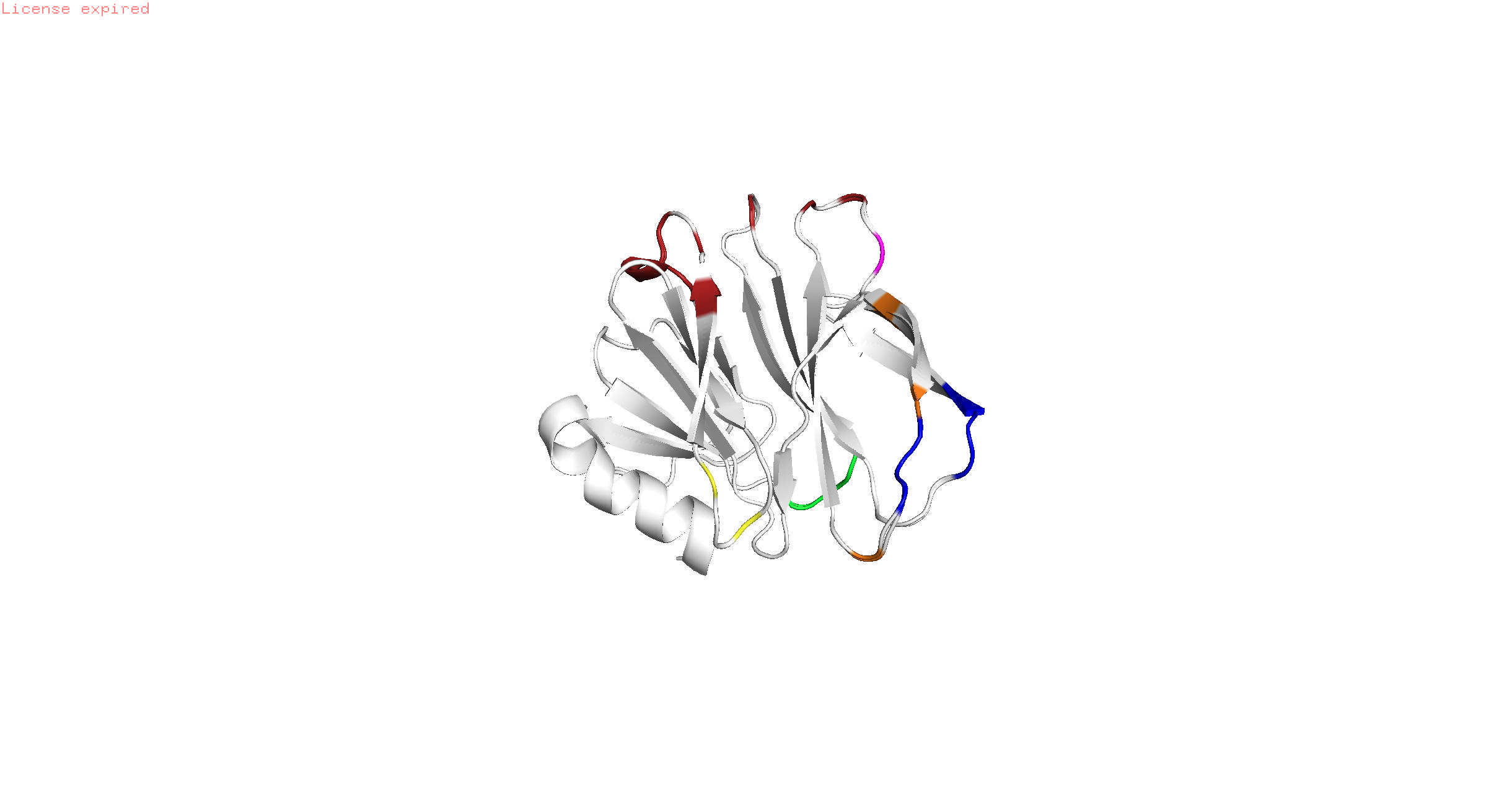

a.

b.

c.

**Supplementary Figure S4.** **Protein model structures comparing the antigenic regions within VP8* of the VP4 protein between Rotarix and the G3P[8] strains**. (a) Perfect alignment of superimposed VP4 structures exhibiting few differences between RV1 and P[8] genotypes associated with G3 rotaviruses in Malawi. VP8*-1 in red, VP8*-2 in blue, VP8*-3 in green and VP8*-4 in yellow. Amino acid position 150 and 131 are colored in magenta and orange respectively. (b and c) surface structural differences between Rotarix and P[8] genotypes associated with rotaviruses in Malawi.
