## Supplementary Table S1 for "Comparative whole genome analysis reveals re-emergence of typical human Wa-like and DS-1-like G3 rotaviruses after Rotarix vaccine introduction in Malawi"

**Supplementary Table S1. Whole genome Nucleotide and amino acid sequence length for Malawian G3 rotavirus strains.**

| **Strain name** | **Sequence-type** | **VP7** | **VP4** | **VP6** | **VP1** | **VP2** | **VP3** | **NSP1** | **NSP2** | **NSP3** | **NSP4** | **NSP5/6** |
| --- | --- | --- | --- | --- | --- | --- | --- | --- | --- | --- | --- | --- |
| **RVA/Human-wt/MWI/BTY22H/2017/G3P[4]** | NT | - | 2359 | 1356 | 3302 | 2684 | 2591 | 1566 | 1059 | 1066 | - | 816 |
|  | AA | - | 775 | 397 | 1088 | 879 | 835 | 493 | 317 | 314 | - | 200 |
| **RVA/Human-wt/MWI/BTY22J/2017/G3P[4]** | NT | 1062 | 2359 | 1356 | 3302 | 2684 | 2591 | 1566 | 1059 | 1066 | 751 | 816 |
|  | AA | 326 | 775 | 397 | 1088 | 879 | 835 | 493 | 317 | 314 | 184 | 200 |
| **RVA/Human-wt/MWI/BTY232/2017/G3P[4]** | NT | 1062 | 2359 | 1356 | 3302 | 2684 | 2591 | 1566 | 1059 | 1066 | 751 | 816 |
|  | AA | 326 | 775 | 397 | 1088 | 879 | 835 | 493 | 317 | 314 | 184 | 200 |
| **RVA/Human-wt/MWI/BTY23I/2018/G3P[4]** | NT | 1062 | 2359 | 1356 | 3302 | 2684 | 2591 | 1566 | 1059 | 1066 | 751 | - |
|  | AA | 326 | 775 | 397 | 1088 | 879 | 835 | 493 | 317 | 314 | 184 | - |
| **RVA/Human-wt/MWI/BTY240/2018/G3P[4]** | NT | 1062 | 2359 | 1356 | 3302 | 2684 | 2591 | 1566 | 1059 | 1066 | 751 | 816 |
|  | AA | 326 | 775 | 397 | 1088 | 879 | 835 | 493 | 317 | 314 | 184 | 200 |
| **RVA/Human-wt/MWI/BTY24R/2018/G3P[4]** | NT | 1062 | 2359 | 1356 | 3302 | 2684 | 2591 | 1566 | 1059 | 1066 | 751 | 816 |
|  | AA | 326 | 775 | 397 | 1088 | 879 | 835 | 493 | 317 | 314 | 184 | 200 |
| **RVA/Human-wt/MWI/BTY250/2018/G3P[4]** | NT | 1062 | 2359 | 1356 | 3302 | 2684 | 2591 | 1566 | 1059 | 1066 | 751 | 816 |
|  | AA | 326 | 775 | 397 | 1088 | 879 | 835 | 493 | 317 | 314 | 184 | 200 |
| **RVA/Human-wt/MWI/BTY25L/2018/G3P[4]** | NT | 1062 | 2359 | 1356 | 3302 | 2684 | 2591 | 1566 | 1059 | 1066 | 751 | 816 |
|  | AA | 326 | 775 | 397 | 1088 | 879 | 835 | 493 | 317 | 314 | 184 | 200 |
| **RVA/Human-wt/MWI/BTY260/2018/G3P[4]** | NT | 1062 | 2359 | 1356 | 3302 | 2684 | 2591 | 1566 | 1059 | 1066 | 751 | 816 |
|  | AA | 326 | 775 | 397 | 1088 | 879 | 835 | 493 | 317 | 314 | 184 | 200 |
| **RVA/Human-wt/MWI/BTY26H/2018/G3P[4]** | NT | 1062 | 2359 | 1356 | 3302 | 2684 | 2591 | 1566 | 1059 | 1066 | 751 | 816 |
|  | AA | 326 | 775 | 397 | 1088 | 879 | 835 | 493 | 317 | 314 | 184 | 200 |
| **RVA/Human-wt/MWI/BTY27C/2018/G3P[4]** | NT | 1062 | 2359 | 1356 | 3302 | 2684 | 2591 | 1566 | 1059 | 1066 | 751 | 816 |
|  | AA | 326 | 775 | 397 | 1088 | 879 | 835 | 493 | 317 | 314 | 184 | 200 |
| **RVA/Human-wt/MWI/BTY27S/2018/G3P[4]** | NT | 1062 | 2359 | 1356 | 3302 | 2684 | 2591 | 1566 | 1059 | 1066 | 751 | 816 |
|  | AA | 326 | 775 | 397 | 1088 | 879 | 835 | 493 | 317 | 314 | 184 | 200 |
| **RVA/Human-wt/MWI/BTY296/2018/G3P[4]** | NT | 1062 | 2359 | 1356 | 3302 | 2684 | 2591 | 1566 | 1059 | 1066 | 751 | 816 |
|  | AA | 326 | 775 | 397 | 1088 | 879 | 835 | 493 | 317 | 314 | 184 | 200 |
| **RVA/Human-wt/MWI/BTY29D/2018/G3P[4]** | NT | 1062 | 2359 | 1356 | 3302 | 2684 | 2591 | 1566 | 1059 | 1066 | 751 | 816 |
|  | AA | 326 | 775 | 397 | 1088 | 879 | 835 | 493 | 317 | 314 | 184 | 200 |
| **RVA/Human-wt/MWI/BTY29E/2018/G3P[4]** | NT | 1062 | 2359 | 1356 | 3302 | 2684 | 2591 | 1566 | 1059 | 1066 | 751 | - |
|  | AA | 326 | 775 | 397 | 1088 | 879 | 835 | 493 | 317 | 314 | 184 | - |
| **RVA/Human-wt/MWI/BTY2A2/2018/G3P[4]** | NT | 1062 | - | 1356 | 3302 | 2684 | 2591 | 1566 | 1059 | 1066 | 751 | - |
|  | AA | 326 | - | 397 | 1088 | 879 | 835 | 493 | 317 | 314 | 184 | - |
| **RVA/Human-wt/MWI/BTY2BG/2018/G3P[4]** | NT | 1062 | 2359 | 1356 | 3302 | 2684 | 2591 | 1566 | 1059 | 1066 | 751 | 816 |
|  | AA | 326 | 775 | 397 | 1088 | 879 | 835 | 493 | 317 | 314 | 184 | 200 |
| **RVA/Human-wt/MWI/BTY2EP/2019/G3P[4]** | NT | 1062 | 2359 | 1356 | 3302 | 2684 | 2591 | 1566 | 1059 | 1066 | 751 | - |
|  | AA | 326 | 775 | 397 | 1088 | 879 | 835 | 493 | 317 | 314 | 184 | - |
| **RVA/Human-wt/MWI/CHX11Q/2019/G3P[4]** | NT | 1062 | 2359 | 1356 | 3302 | 2684 | 2591 | 1566 | 1059 | 1066 | 751 | 816 |
|  | AA | 326 | 775 | 397 | 1088 | 879 | 835 | 493 | 317 | 314 | 184 | 200 |
| **RVA/Human-wt/MWI/CHX11X/2019/G3P[4]** | NT | 1062 | 2359 | 1356 | 3302 | 2684 | 2591 | 1566 | 1059 | 1066 | 751 | - |
|  | AA | 326 | 775 | 397 | 1088 | 879 | 835 | 493 | 317 | 314 | 184 | 200 |
| **RVA/Human-wt/MWI/BTY2CM/2018/G3P[6]** | NT | 1062 | 2359 | 1356 | 3302 | 2684 | 2591 | 1566 | 1059 | 1066 | 751 | 816 |
|  | AA | 326 | 775 | 397 | 1088 | 879 | 835 | 493 | 317 | 314 | 184 | 200 |
| **RVA/Human-wt/MWI/CBTY2BD/2018/G3P[8]** | NT | 1062 | 2359 | 1356 | 3302 | 2729 | 2591 | 1567 | 1059 | 1074 | 750 | 664 |
|  | AA | 326 | 775 | 397 | 1088 | 894 | 835 | 493 | 317 | 314 | 184 | 197 |
| **RVA/Human-wt/MWI/BTY2EM/2019/G3P[8]** | NT | 1062 | 2359 | 1356 | 3302 | 2729 | 2591 | 1567 | 1059 | 1074 | 750 | 664 |
|  | AA | 326 | 775 | 397 | 1088 | 894 | 835 | 493 | 317 | 314 | 184 | 197 |
| **RVA/Human-wt/MWI/BTY2GA/2019/G3P[8]** | NT | 1062 | 2359 | 1356 | 3302 | 2729 | 2591 | 1567 | 1059 | 1074 | 750 | 664 |
|  | AA | 326 | 775 | 397 | 1088 | 894 | 835 | 493 | 317 | 314 | 184 | 197 |
| **RVA/Human-wt/MWI/BTY2GC/2019/G3P[8]** | NT | 1062 | 2359 | 1356 | 3302 | 2729 | 2591 | 1567 | 1059 | 1074 | 750 | 664 |
|  | AA | 326 | 775 | 397 | 1088 | 894 | 835 | 493 | 317 | 314 | 184 | 197 |
| **RVA/Human-wt/MWI/CHX11U/2019/G3P[8]** | NT | 1062 | 2359 | 1356 | 3302 | 2729 | 2591 | 1567 | 1059 | 1074 | 750 | 664 |
|  | AA | 326 | 775 | 397 | 1088 | 894 | 835 | 493 | 317 | 314 | 184 | 197 |
| **RVA/Human-wt/MWI/CHX11S/2019/G3P[8]** | NT | 1062 | 2359 | 1356 | 3302 | 2729 | 2591 | 1567 | 1059 | 1074 | 750 | 664 |
|  | AA | 326 | 775 | 397 | 1088 | 894 | 835 | 493 | 317 | 314 | 184 | 197 |
