## Supplementary Table S2 for "Comparative whole genome analysis reveals re-emergence of typical human Wa-like and DS-1-like G3 rotaviruses after Rotarix vaccine introduction in Malawi"

**Supplementary Table S2**: **Lineages for Wa-like (green) and DS-1-like (red) genome segments associated with re-emergent G3 rotavirus strains in Malawi**. There is no lineage framework available for Wa-like genome segments hence the only available Wa-like reference sequences that were assigned lineages previously were utilised to assign lineages to VP4 and VP7 encoding genome segments of the Malawian G3 strains.

|  | **VP7** | **VP4** | | **VP6** | **VP1** | **VP2** | **VP3** | | **NSP1** | **NSP2** | **NSP3** | **NSP4** | **NSP5** |
| --- | --- | --- | --- | --- | --- | --- | --- | --- | --- | --- | --- | --- | --- |
| **G3P[4]** | III | IVa | V | V | V | IVa | VII | VI | IVa | V | V | VI | IVa |
| **G3P[6]** | III | Ia | | V | V | IVa | VII | | IVa | V | V | VI | IVa |
| **G3P[8]** | III | III | | - | - | - | - | | - | - | - | - | - |
