## Supplementary Table S3 for "Comparative whole genome analysis reveals re-emergence of typical human Wa-like and DS-1-like G3 rotaviruses after Rotarix vaccine introduction in Malawi"

**Supplementary Table S3. Alignment of antigenic residues of VP4 protein between the P[8] component of Rotarix and VP4 component of Malawian G3 strains**. Antigenic residues are divided into four antigenic epitopes for VP8* (8-1, 8-2, 8-3 and 8-4) and five antigenic epitopes for VP5* (5-1, 5-2, 5-3, 5-4, and 5-5). Amino acid changes that have been shown to escape neutralisation with monoclonal antibodies are indicated with a black dot

|  |  |  | **8*-1** | | | | | | | | | | |  | **8*-2** | |  | **8*-3** | | | | | | | | |  | **8*-4** | | |  | **5*-1** | | | | | | | |  | **5*-2** |  | **5*-3** |  | **5*-4** |  | **5*-5** |
| --- | --- | --- | --- | --- | --- | --- | --- | --- | --- | --- | --- | --- | --- | --- | --- | --- | --- | --- | --- | --- | --- | --- | --- | --- | --- | --- | --- | --- | --- | --- | --- | --- | --- | --- | --- | --- | --- | --- | --- | --- | --- | --- | --- | --- | --- | --- | --- |
|  |  |  | **●** | **●** | **●** | **●** | **●** | **●** |  |  | **●** |  |  |  | **●** | **●** |  |  | **●** |  | **●** |  |  | **●** | **●** | **●** |  | **●** | **●** | **●** |  | **●** | **●** | **●** | **●** | **●** | **●** | **●** | **●** |  | **●** |  | **●** |  | **●** |  | **●** |
| **Strain** | **Genotype** | **Lineage** | **100** | **146** | **148** | **150** | **188** | **190** | **192** | **193** | **194** | **195** | **196** |  | **180** | **183** |  | **113** | **114** | **115** | **116** | **125** | **131** | **132** | **133** | **135** |  | **87** | **88** | **89** |  | **384** | **386** | **388** | **393** | **394** | **398** | **440** | **441** |  | **434** |  | **459** |  | **429** |  | **306** |
| **ROTARIX** | P[8] | I | D | S | Q | E | S | T | N | L | N | N | I |  | T | A |  | N | P | V | D | S | S | N | D | N |  | N | T | N |  | Y | F | I | W | P | G | R | T |  | P |  | E |  | L |  | L |
| **BTY2GC** | P[8] | III | * | * | * | D | * | * | * | * | * | * | * |  | * | * |  | * | * | * | * | N | R | * | * | D |  | * | * | * |  | * | * | * | * | * | * | * | * |  | * |  | * |  | * |  | * |
| **BTY2GA** | P[8] | III | * | * | * | D | * | * | * | * | * | * | * |  | * | * |  | * | * | * | * | N | R | * | * | D |  | * | * | * |  | * | * | * | * | * | * | * | * |  | * |  | * |  | * |  | * |
| **BTY2BD** | P[8] | III | * | * | * | D | * | * | * | * | * | * | * |  | * | * |  | * | * | * | * | N | R | * | * | D |  | * | * | * |  | * | * | * | * | * | * | * | * |  | * |  | * |  | * |  | * |
| **BTY2EM** | P[8] | III | * | * | * | D | * | * | * | * | * | * | * |  | * | * |  | * | * | * | * | N | R | * | * | D |  | * | * | * |  | * | * | * | * | * | * | * | * |  | * |  | * |  | * |  | * |
| **CHX11U** | P[8] | III | * | * | * | D | * | * | * | * | * | * | * |  | * | * |  | * | * | * | * | N | R | * | * | D |  | * | * | * |  | * | * | * | * | * | * | * | * |  | * |  | * |  | * |  | * |
| **CHX11S** | P[8] | III | * | * | * | D | * | * | * | * | * | * | * |  | * | * |  | * | * | * | * | N | R | * | * | D |  | * | * | * |  | * | * | * | * | * | * | * | * |  | * |  | * |  | * |  | * |
| **BTY2EP** | P[4] | IVa | * | * | * | D | * | * | D | * | * | N | * |  | * | * |  | S | * | T | N | N | E | * | S | D |  | * | * | D |  | * | * | L | * | * | * | * | * |  | * |  | * |  | * |  | * |
| **BTY2BG** | P[4] | IVa | * | * | * | D | * | * | D | * | * | N | * |  | * | * |  | S | * | T | N | N | E | * | S | D |  | * | * | D |  | * | * | L | * | * | * | * | * |  | * |  | * |  | * |  | * |
| **BTY29D** | P[4] | V | * | * | * | D | * | * | D | * | * | N | * |  | * | * |  | S | Q | T | N | N | E | * | S | D |  | * | A | D |  | * | * | L | * | * | * | * | * |  | * |  | * |  | * |  | * |
| **BTY22H** | P[4] | V | * | * | * | D | * | * | D | * | * | N | * |  | * | * |  | S | Q | T | N | N | E | * | S | D |  | * | * | D |  | * | * | L | * | * | * | * | * |  | * |  | * |  | * |  | * |
| **BTY2CM** | P[6] | Ia | * | * | S | * | * | * | * | * | S | * | V |  | * | * |  | T | N | Q | S | T | E | * | N | S |  | T | N | Q |  | * | * | * | * | * | * | * | * |  | * |  | * |  | * |  | * |
